# Streamlined CRISPR-based assays for detection and subtyping of avian influenza

**DOI:** 10.1101/2025.08.21.25334105

**Authors:** Yujia Huang, Andrew Guo, Gordon Adams, Jacob E. Lemieux, Cameron Myhrvold

## Abstract

Avian influenza viruses (AIVs) are zoonotic pathogens that pose an increasing global threat due to their potential for significant economic losses in agriculture, spillover into humans, and the risk of a pandemic should human-to-human transmission occur. These concerns underscore the need for rapid, sensitive and specific tools to detect and differentiate circulating AIV subtypes and clades. Current AIV diagnostic methods rely on specialized equipment and trained personnel, limiting their use in the field and in low-resource settings. Here, we extended SHINE (Streamlined Highlighting of Infections to Navigate Epidemics), a CRISPR-based platform, to detect and subtype AIVs. We designed, optimized, and validated SHINE assay for the H5 AIV detection using both fluorescence and lateral flow readout, achieving 100% specificity with PCR-based assays when tested on seasonal influenza-positive clinical samples, and a limit of detection of 121.7 copies/μL on vaccine-derived H5 viral seedstocks. To expand the scope of avian influenza detection, we also designed and validated a SHINE assay targeting the 2.3.4.4b A(H5N1) lineage, in response to the ongoing H5N1 outbreak in cattle in the United States, and a SHINE assay specific to Eurasian H7 lineage to discriminate against North American H7 lineage. Together, these SHINE assays offer a promising platform for AIV diagnosis and surveillance, particularly in settings with limited laboratory infrastructure.

## Introduction

Avian influenza is a zoonotic disease caused by avian influenza A viruses (AIVs) that poses a significant threat to both animal and human health (*1*). These viruses are classified into subtypes with varying host range and pathogenicity based on two distinct surface glycoproteins: hemagglutinin (H1-H16), and neuraminidase (N1-N9) (*2*). Based on their pathogenicity, AIVs are further classified into low pathogenic avian influenza (LPAI) and highly pathogenic avian influenza (HPAI), with HPAI subtypes such as H5N1 and H7N9 associated with significantly higher mortality in both birds and humans (*3–5*). Notable outbreaks, such as the H5N1 outbreak in Hong Kong in 1997 and the H7N9 outbreak in China in 2013, demonstrated that HPAI can cause severe diseases with high case fatality rates of at least 30% (*6–10*).

Recent HPAI outbreaks have further highlighted the need for robust technologies for both AIV surveillance and diagnosis. Since its first detection in January 2022, HPAI H5 has infected more than 13,000 wild birds and 173,000,000 poultry, resulting in economic losses ranging from $14 billion to $164 billion (*11*, *12*). In March 2024, an unprecedented clade 2.3.4.4b A(H5N1) outbreak in dairy cattle was reported in Texas, marking a significant shift in hosts for AIVs (*13*, *14*). To date, this virus has spread across 17 US states, infecting over 1,000 cattle herds and showing sustained mammalian transmission with 70 human cases reported as of June 3rd, 2025 (*15*). As novel, highly pathogenic AIVs may continue to emerge, there remains a critical need for rapid, subtype- and clade-specific diagnostic tools to support surveillance, public health preparedness, and outbreak response.

Current avian influenza diagnosis primarily relies on laboratory-based testing using virus isolation, immunoassays, RT-PCR, and sequencing methods. Virus isolation is considered the gold standard for diagnosing AIV, but is restricted to biosafety level-3 laboratories and requires highly trained personnel (*16*). In contrast, immunoassays can offer rapid, point-of-care diagnosis within approximately 15 minutes, but exhibit reduced sensitivity and are not specific enough for clade-specific discrimination (*17–19*). Nucleic acid detection methods like RT-PCR have been widely adopted in both outbreak response and wild bird surveillance due to their high sensitivity and adaptability (*20*, *21*). However, PCR-based methods still require thermocycling equipment and trained personnel, limiting their deployment in field settings. Next-generation sequencing (NGS), which typically relies on PCR for target enrichment, enables nucleotide-level resolution, but remains limited for routine clinical use due to its high cost and long turnaround time (*22–24*). Consequently, current diagnostic methods for AIV detection require substantial laboratory equipment and infrastructure or suffer from reduced sensitivity and specificity that hamper the public response to AIV epidemic spread and host spillover events.

Recently developed CRISPR-based diagnostic assays (CRISPR-Dx) offer a promising approach for detecting and identifying AIV subtypes. CRISPR-Dx are highly specific, sensitive, and programmable, relying on complementary base pairing between a CRISPR RNA (crRNA) and the target of interest to activate the nuclease activity of CRISPR-associated enzymes such as Cas12 or Cas13, resulting in a fluorescent or colorimetric signal through reporter cleavage (*25*, *26*). Several groups have previously explored CRISPR-Dx for avian influenza detection, pairing Cas12a or Cas13a detection with pre-amplification steps to enhance sensitivity (Supplementary Table 1) (*27–33*). However, all of these methods involve multi-step workflows that separate amplification and detection steps, which increases the risk of cross-contamination and limits their applicability for clinical testing. Therefore, there is an unmet need for simple, sensitive and CRISPR-Dx methods that enable surveillance for AIV subtyping at the point of care and point of need.

Here, we develop SHINE (Streamlined Highlighting of Infections to Navigate Epidemics) (*34–36*) assays for detecting and differentiating between AIVs using both fluorescence and lateral flow readouts. We demonstrate that SHINE can achieve highly sensitive and specific detection against multiple H5 strains at different levels of genetic resolution, ranging from broad subtype-level detection to fine clade-specific discrimination. We applied multiple optimization strategies to the H5 SHINE assay and comprehensively characterized its analytical sensitivity, limit of detection, and specificity. We also developed a SHINE assay against the H7 Eurasian lineage to expand the scope of AIV diagnostics. Together, these SHINE assays present a promising approach for rapid, low-cost, and accessible testing that can support the detection and surveillance of avian influenza, particularly in settings with limited laboratory infrastructure.

## Results

### Development of a prototype H5 assay using SHINE

We sought to develop prototype CRISPR-Dx assays for AIVs, focusing on the detection of H5 AIVs. To improve field deployability, we used the SHINE platform, a one-pot CRISPR-Dx technology optimized for point-of-need use. In this SHINE workflow, avian influenza genomes are reverse transcribed and amplified by SSIV reverse transcriptase, RPA, and T7 RNA polymerase. Complementary base pairing between a Cas13-crRNA complex and the amplified genome of avian flu can activate the *trans* cleavage activity of Cas13, cleaving RNA-based reporters and leading to a fluorescent or colorimetric signal (Figure 1A).

**Figure 1.**
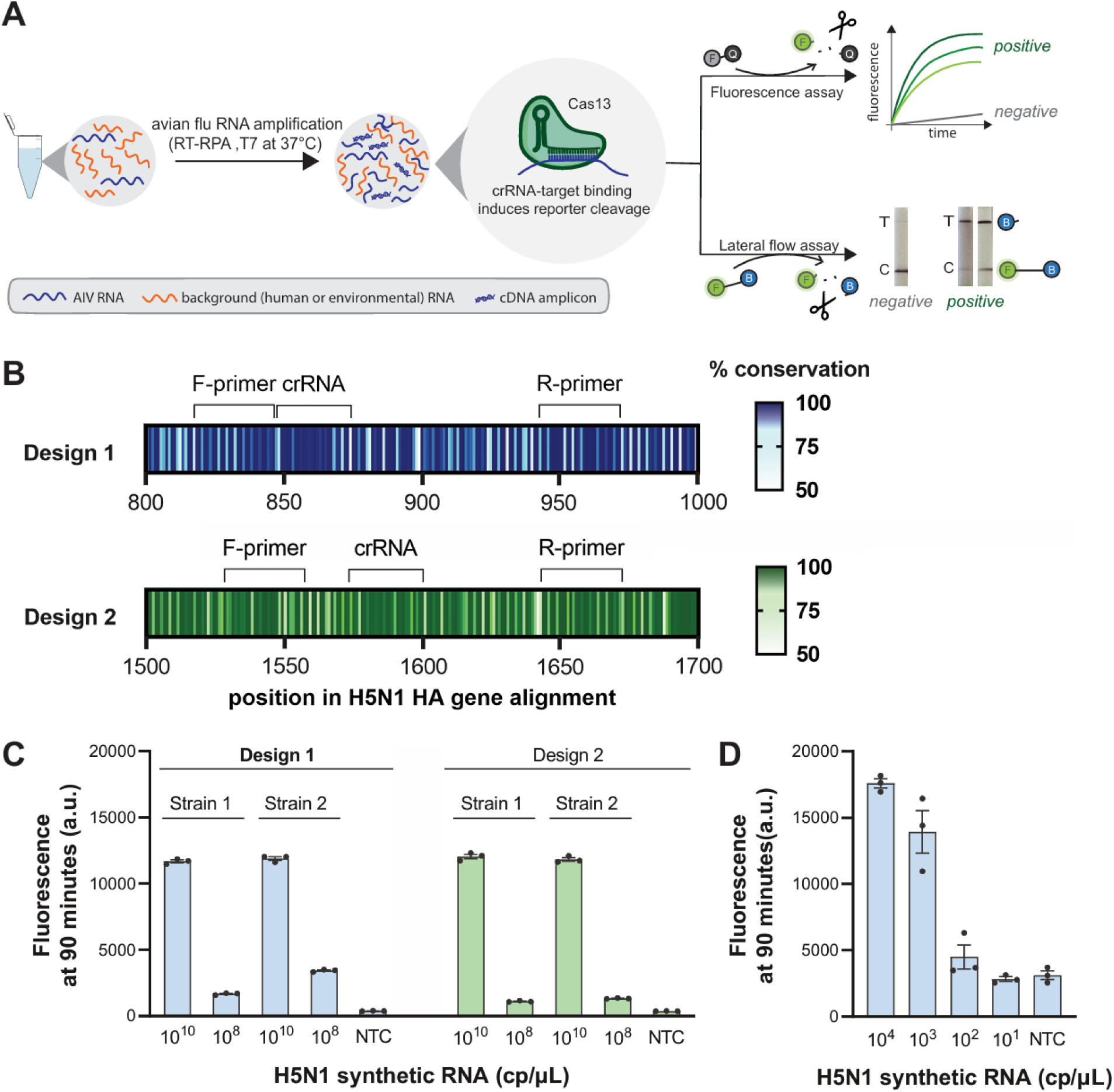
Initial development of the prototype H5 SHINE assay. **A)** Schematic of avian flu SHINE workflow. RNA undergoes single-step amplification, consisting of reverse-transcription recombinase polymerase amplification (RT-RPA), T7 transcription, and Cas13 detection in a single tube. Fluorescence readout is achieved through Cas13-mediated cleavage of a FAM quenched fluorescent reporter (F-Q) or paper-based readout through cleavage of a FAM-biotin (F-B) reporter. For paper-based readouts, positive samples exhibit a test (T) band, sometimes accompanied by a control (C) band at lower target concentrations. Negative samples show only a strong C band. **B)** Two designs targeting different regions of the H5N1 hemagglutinin (HA) gene are shown. Heatmaps show local sequence conservation across aligned H5N1 HA gene sequences, with annotated regions for the forward primer (F-primer), crRNA, and reverse primer (R-primer). The color gradient is non-linear, regions with >85% conservation (darker colour) are considered highly conserved and were prioritized. **C)** Cas13a detecting synthetic RNA for two assay designs against two H5N1 strains (Strain 1: A/white-tailed_eagle/Estonia/ TA2111864-2_21VIR7512-6/2021, Strain 2: A/mute_swan/England/328136/2022). Design 1 (bolded) was selected for further development. **D)** Fluorescence readout of H5N1 SHINE assays on a serial dilution of H5N1 synthetic RNA. For **C** and **D**, values are mean ± standard deviation of 3 technical replicates after 90 minutes. NTC: no target control.

To design the sequences of crRNAs and RPA primers for our H5 SHINE assay, we used ADAPT (Activity-informed Design with All-inclusive Patrolling of Targets), a software platform that leverages machine learning to maximize sensitivity and target specificity in Cas13-based assay design (*37*). We focused our designs on the H5N1 influenza genome segment that encodes for the hemagglutinin (HA) protein, as this segment is unique to H5 viruses. We selected two highest-scoring primer-crRNA sets, with a predicted coverage of 98.72% and 96.18%, respectively, for experimental validation (Figure 1B). We validated crRNA cleavage activity with high concentrations of synthetic RNA input from two H5N1 strains, A/white-tailed eagle/Estonia/1945381/2021 (H5N1) and A/mute swan/England/328136/2022 (H5N1) (Figure 1C, Supplementary Figure 1A). Design 1 (crRNA targeting nucleotides 847-874 of the consensus sequence of the H5 segment) showed higher activity with 10^8^ copies of target input for both targets, so we selected this design for subsequent H5 SHINE assay development.

To evaluate the performance of integrating isothermal amplification with Cas13 detection, we tested the SHINE assay using synthetic nucleic acid inputs. We employed two types of targets: double-stranded DNA templates that encoded the entire H5 segment to assess amplification and detection capability, and *in vitro* transcribed H5 segment RNA templates. The SHINE assay demonstrated robust sensitivity with DNA input, enabling consistent detection across all concentrations tested, ranging from 10^4^ to 10^1^ copies/μL (Supplementary Figure 1B, C). However, sensitivity was substantially reduced with RNA inputs, only achieving consistent detection at 10^3^ copies/μL, likely due to competition between reverse transcription and Cas13 detection (Figure 1D, Supplementary Figure 1D). A concentration-dependent reduction in fluorescence was observed, suggesting that signal strength is proportional to input concentration. These results demonstrate that the prototype SHINE assay we designed has the potential to be used for H5N1 surveillance with further optimization to improve sensitivity and field applicability.

### Optimization of the H5 SHINE assay

To improve the sensitivity and reduce the turnaround time of the prototype H5 SHINE assay, we aimed to achieve reliable detection of RNA synthetic targets at 10^2^ copies/μL within 60 minutes. We sought to improve reaction kinetics by reducing inter-enzymatic interference and boosting reaction rate for individual enzymes. A previous study demonstrated that incorporating a non-T7 primer can enhance amplification in the early stage, likely by reducing competition between amplification and detection, thereby improving sensitivity at early time points (*38*). We evaluated a range of T7-primer to non-T7-primer ratios and identified that a 1:3 ratio works best for our prototype H5 assay (Figure 2A, Supplementary Figure 2A, 3A). Additionally, we tested conditions with increased RPA primer concentrations and found that a slightly higher RPA primer concentration can enhance amplification dynamics, improving assay sensitivity (Supplementary Figure 2B, 3B). Subsequent testing was performed with individually altered reporter, RNase H, and magnesium concentrations (Supplementary Figure 2C, D, E, 3C, D, E). These optimizations, when combined, yielded an optimized H5 assay (hereafter referred to as SHINE-H5) that outperformed the prototype assay, achieving successful detection for all tested target concentrations from the NTC within 60 minutes (Figure 2B, Supplementary Figure 4).

**Figure 2.**
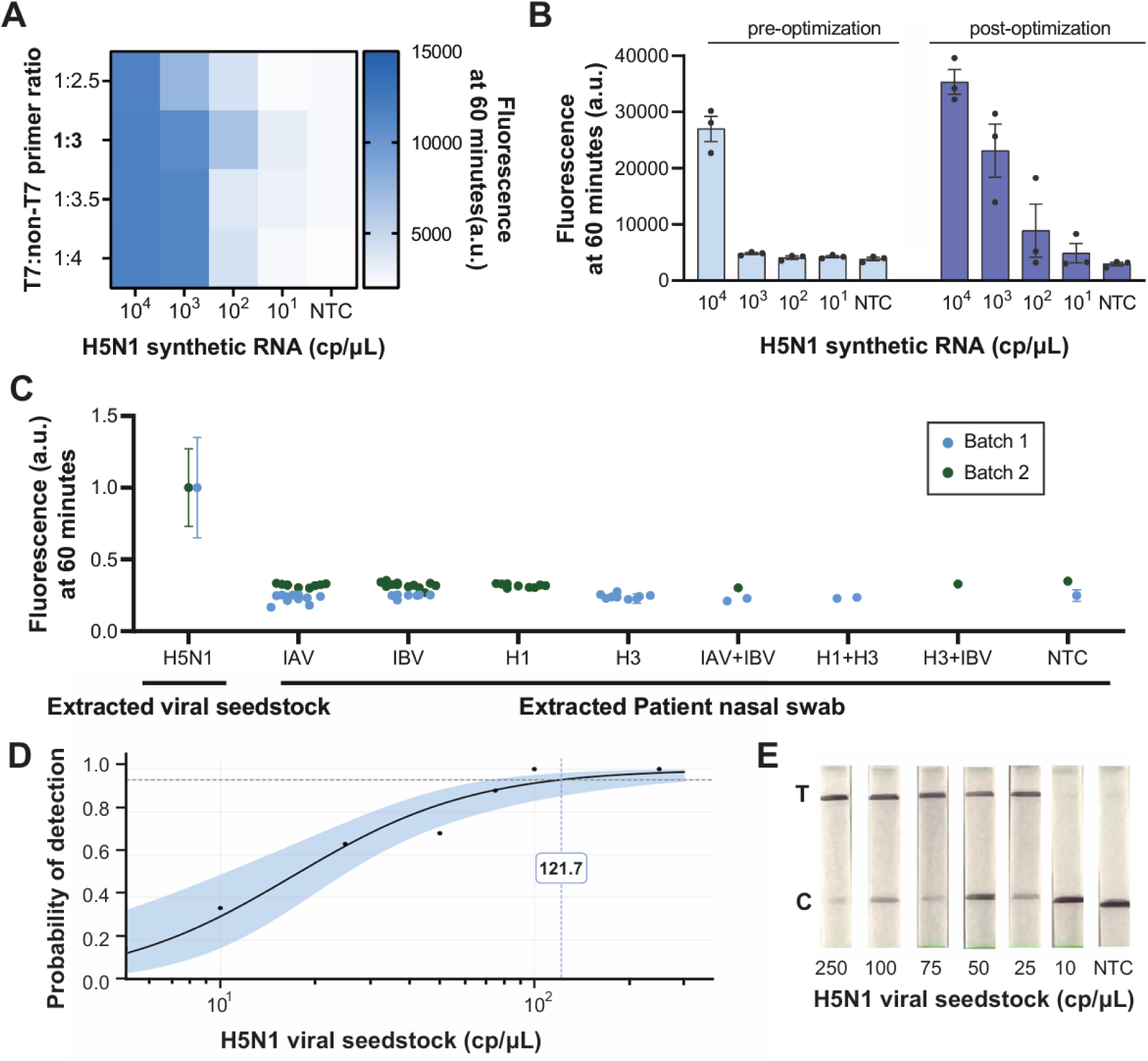
Optimization and detailed characterization of the H5 SHINE assay. **A)** Heatmap showing the H5 SHINE assay performance at conditions with different non-T7 primer : T7-primer ratios **B)** Fluorescence readout of pre- and post-optimization SHINE assays on a serial dilution of H5N1 synthetic RNA. Values are mean fluorescence ± standard deviation of 3 technical replicates after 60 minutes. **C)** Normalized fluorescence readout of optimized SHINE-H5 on RNA extracted from H5N1 viral seedstock (10^2^ cp/μL) and from clinical NP swabs characterized by NAAT (BioFire) or qPCR. Fluorescence values were normalized to the H5N1 positive control for each sample set. Each dot shows mean ± standard deviation of 3 technical replicates of a specific patient sample after 60 minutes. See Supplementary Table 3 for detailed patient information and qPCR/NAAT/SHINE readout. **D)** Limit of detection (LoD) for H5N1 viral seedstock. The black curve shows a fitted logistic regression model, and the shaded region represents 95% Wald confidence interval. LOD = 121.7 (63.14 – 234.55) cp/μL, N=20 technical replicates were used at each dilution. **E)** Paper-based colorimetric readout of H5N1 SHINE assay on a serial dilution of RNA extracted from H5N1 viral seedstocks after 60-minute incubation. NTC: no target control.

To assess the specificity of SHINE-H5, we first evaluated its performance against six different vaccine-derived viral seedstocks. SHINE-H5 demonstrated high target specificity, producing high fluorescence signals against the desired target and negligible fluorescence for non-target viruses (Supplementary Figure 5A, Supplementary Table 2). Given that the clinical syndrome avian influenza in humans overlaps with several common respiratory illnesses (*39*), we next evaluated the clinical specificity of SHINE-H5 using nasopharyngeal (NP) swab specimens. Since H5N1 infections in humans are exceedingly rare, seasonal influenza viruses represent the most clinically relevant and high-prevalence specimen type for cross-reactivity testing (*40*, *41*). A total of 64 NP swab specimens with positive clinical influenza results—either influenza A with H-subtype information, influenza A without subtype information, or influenza B (Supplementary Table 3)—were extracted and tested with the SHINE assay. All specimens were negative for avian H5 by qPCR as described in Methods. SHINE-H5 showed 100% (64/64) specificity across two clinical sample sets, covering both single and co-infections with both H1N1 and H3N2 seasonal influenza viruses (Figure 2C, Supplementary Figure 5B, C).

To further evaluate the analytical performance of SHINE-H5, we determined its limit of detection (LoD) using serial dilutions of H5N1 viral seedstock in viral transport media, ranging from 10 to 250 copies/μL, as input to SHINE-H5. LoD fluorescence cutoffs were established as the mean fluorescence of the extraction controls (viral transport media alone) plus three times their standard deviation (SD). The LoD was defined as the concentration at which ≥95% (19/20) of the SHINE replicates exceeded this threshold, as determined by a logistic regression analysis. The LoD for H5N1 viral seedstock was calculated to be 121.7 (n positive/N total, 63.14 – 234.55, 95% CI) copies/μL (Figure 2D, Supplementary Figure 6).

To enable avian influenza surveillance in PON settings such as cattle or poultry farms, we adapted SHINE-H5 for a lateral flow readout format. In this adapted assay, the FAM quenched reporter is replaced by a biotin-FAM reporter. Uncleaved reporters are captured at the lower streptavidin band while cleaved reporters migrate to the upper test band, so the appearance of the upper band is interpreted as a positive result. With a larger (40μL) reaction volume, the lateral flow version of SHINE-H5 (SHINE-H5-LFA) was able to detect H5N1 viral seedstock down to 25 copies/μL (Figure 2E). SHINE-H5-LFA also demonstrated strong specificity like its fluorometric counterpart, as confirmed by testing a subset of 14 patient samples positive for seasonal influenza but negative for H5 (Supplementary Figure 7, Supplementary Table 3).

### Design and validation of SHINE assay targeting clade 2.3.4.4b A(H5N1)

In addition to developing a SHINE assay for detection of all H5 AIVs, we developed a clade-specific SHINE assay specifically targeting clade 2.3.4.4b A(H5N1), which has been dominant in global H5 phylogeny since 2021 based on available sequences in GISAID (Figure. 3A) and is associated with the ongoing H5N1 outbreak in cattle in the United States. As of June 5, 2025, this outbreak has affected 1,073 dairy herds nationwide, underscoring the urgent need for rapid, clade-specific detection tools (*42*). We curated an on-target dataset comprising clade 2.3.4.4b A(H5N1) sequences and an off-target consisting of non-2.3.4.4b H5 sequences from NCBI Influenza Virus and GISAID EpiFlu databases. To improve the specificity of this assay, we employed BADGERS (Building Artificial Diagnostic Guides by Exploring Regions of Sequences), which builds on the machine learning methods of ADAPT by integrating additional search algorithms that explore the fitness landscape of different crRNA assays (*43*). BADGERS has been shown to significantly outperform ADAPT for sequence variant identification down to the single-nucleotide resolution, which enables discrimination of clade and non-clade H5 sequences. Using BADGERS, we designed three clade-specific crRNA candidates (Figure 3B) and manually adjusted RPA primers from SHINE-H5, shifting forward primer four nucleotides upstream to introduce an additional mismatch that enhances target discrimination while preserving high sequence conservation within the 2.3.4.4b H5 clade.

**Figure 3.**
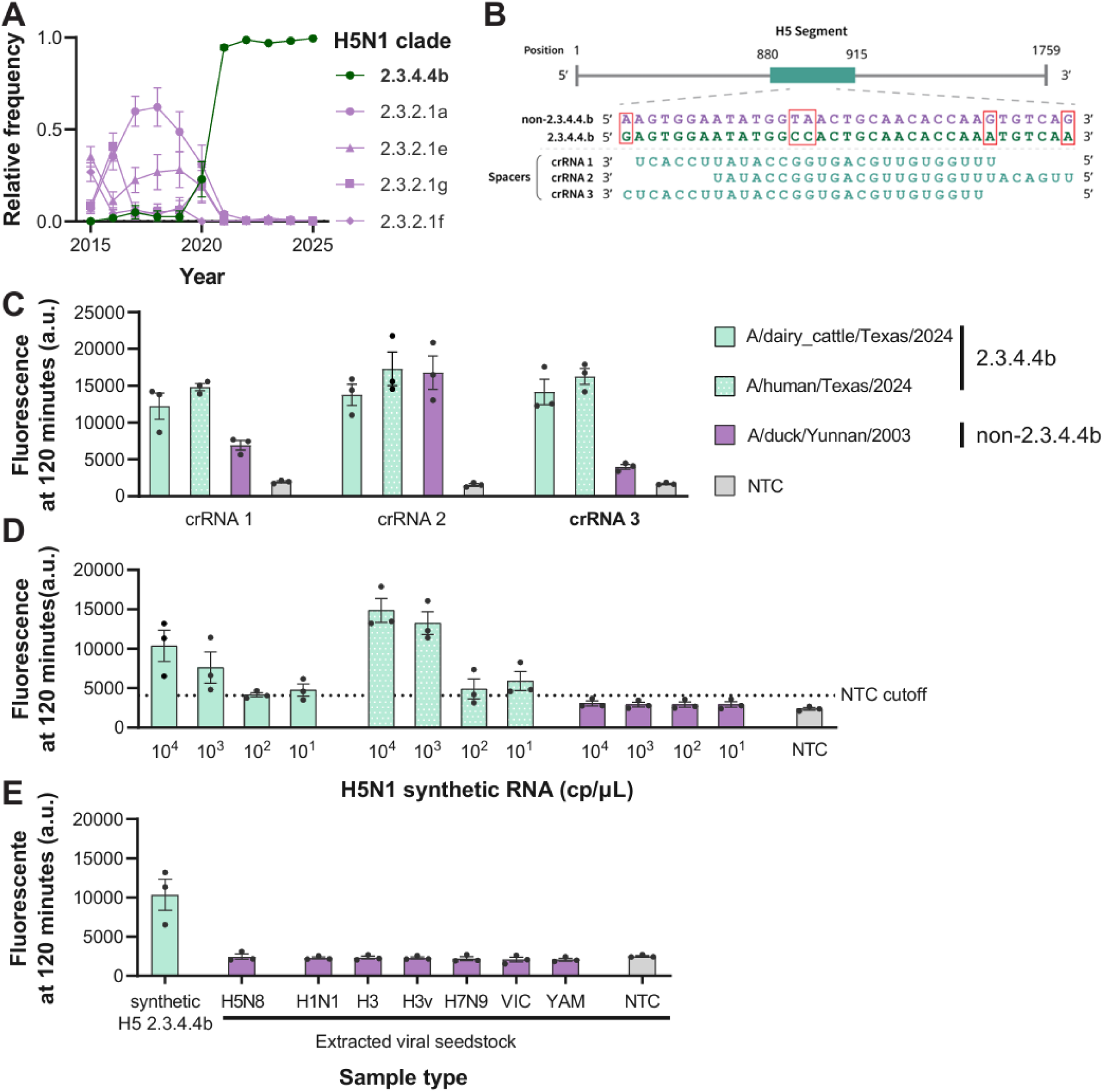
Development of clade 2.3.4.4b A(H5N1) SHINE assay. **A)** Yearly relative frequencies of the five most common H5 clades between 2015-2025. H5 sequences from North America and Eurasia were obtained from GISAID. **B)** Alignment of crRNA sequence and representative H5 gene sequences from clade 2.3.4.4b and non-2.3.4.4b strains at the candidate crRNA binding site. The designed crRNAs show perfect reverse complementarity to the 2.3.4.4b target sequence and multiple mismatches with non-2.3.4.4b sequences, highlighting the potential for selectively detecting 2.3.4.4b H5 viruses. **C)** Cas13a detecting synthetic targets for three different crRNA designs against two different on-targets and one off-target at both 10^10^ copies/μL. **D)** Fluorescence readout of 2.3.4.4b clade-specific H5N1 SHINE assays on a serial dilution of 2.3.4.4b and non-2.3.4.4b H5N1 synthetic RNA, cutoff is mean fluorescence of NTC + 6×standard deviation. **E)** Cross reactivity panel for 2.3.4.4b clade-specific H5N1 SHINE assay against various seasonal influenza viral seedstocks. See Supplementary Table 2 for strain information. For **C,D,E,** values are mean fluorescence ± standard deviation of 3 technical replicates after 120 minutes. NTC: no target control.

We then selected three representative H5N1 viral sequences to evaluate the specificity of the three crRNA candidates. H5 sequences from the isolates A/cattle/Texas/56283/2024 and A/human/Texas/37/2024, both from the ongoing H5N1 outbreak in Texas, were chosen as representative sequences for the clade 2.3.4.4b A(H5N1). Additionally, H5 sequence from isolate A/duck/Yunnan/215/2003 (H5N1) was selected as a non-2.3.4.4.b H5 sequence as an off-target control. Among the three crRNA candidates, crRNA3 (targeting consensus sequence of H5 segment from clade 2.3.4.4b, starting at nucleotide position 880) was selected for further 2.3.4.4b clade-specific H5N1 SHINE assay (hereafter referred to as SHINE-H5-CS) development due to its high on-target activity and minimal off-target activity (Figure 3C, Supplementary Figure 8).

To enhance assay specificity and enable detection at lower viral loads, we designed 2.3.4.4b clade-specific RPA primers and integrated them with crRNA 3 to develop SHINE-H5-CS. SHINE-H5-CS consistently detected both 2.3.4.4b H5 target RNA across all tested concentrations higher than 10^2^ copies/μL while showing minimal cross-reactivity with non-2.3.4.4b H5 target (Figure 3C, D Supplementary Figure 9). These results demonstrate that SHINE-H5-CS reliably discriminates between 2.3.4.4b and non-2.3.4.4b H5 sequences across a wide range of input concentrations.

To further evaluate the specificity of SHINE-H5-CS, we assembled a cross-reactivity panel including various seasonal influenza viral seedstocks covering strains from both Influenza A and B viruses. Testing the assays using this panel demonstrated high specificity, with high fluorescence against the desired target and negligible fluorescence against non-preferred targets (Figure 3E, Supplementary Table 2). SHINE-H5-CS exhibited minimal off-target activity and effectively distinguished the clade 2.3.4.4b A(H5N1) from seasonal influenza viruses.

### A lineage-specific SHINE assay detects Eurasian H7 influenza

While multiple influenza subtypes circulate in domestic poultry, the H5 and H7 subtypes are of particular concern because of their potential to evolve into highly pathogenic strains. Phylogenetic analysis of the HA gene from 25 representative H7 strains revealed clear separation into two major lineages, Eurasian and North American (Figure 4A), which reflected the geographic distribution of their avian hosts, aligning with respective migratory bird flyways (*44*). Eurasian H7 strains share a similar α-2-3 sialic acid receptor binding preference with recent HPAI H5N1 viruses, indicating a higher risk of human infection. Past Eurasian H7 AIV outbreaks such as the 2013 H7N9 epidemic that caused over 1,500 human infections with a fatality rate of around 40% (*45*, *46*) underscore the serious public health threat posed by this subtype. Given the global public health relevance and widespread circulation of Eurasian H7 AIVs, we prioritized SHINE assay development for this lineage to expand the scope of avian influenza detection.

**Figure 4.**
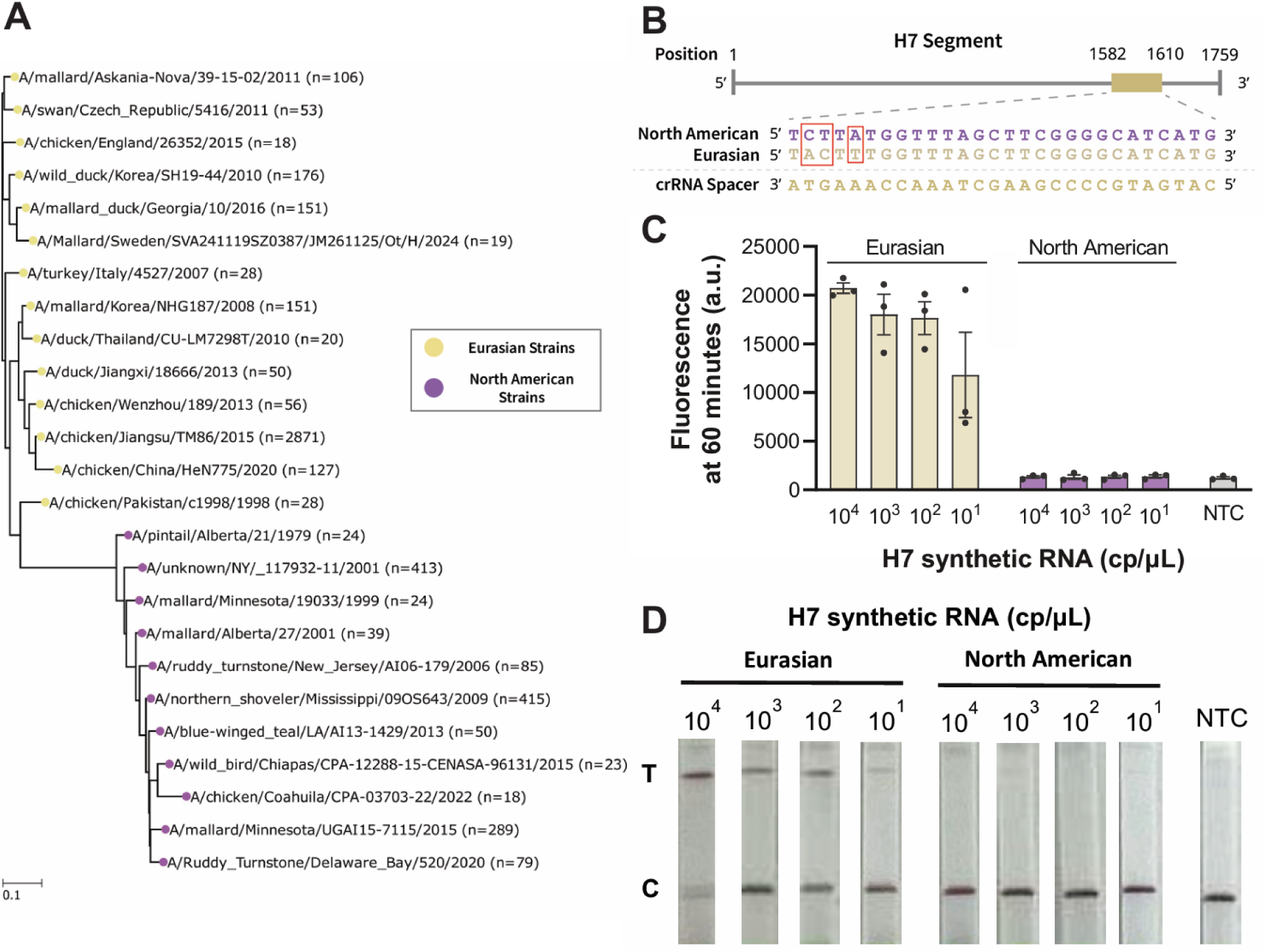
Development of SHINE-H7-Eurasian. **A)** Phylogenetic tree of the H7 subtype hemagglutinin gene. All Eurasian and North American H7 sequences were collected from the GISAID EpiFlu database. Representative sequences were identified via clustering methods. Phylogenetic analysis of these sequences shows that H7 diverges into Eurasian and North American H7 lineages with minimal overlap. **B)** Alignment of crRNA sequence and consensus H7 gene sequences from Eurasian and North American lineages at the crRNA binding site. The designed crRNAs show perfect reverse complementarity to the Eurasian lineage sequence and multiple mismatches with American lineage sequences, highlighting the potential for selectively differentiating these lineages. **C)** Fluorescence readout of H7 Eurasian SHINE assays on a serial dilution of Eurasian and American H7 synthetic RNA. Values are mean fluorescence ± standard deviation of 3 technical replicates after 60 minutes. **D)** Paper-based colorimetric readout of H7 Eurasian SHINE assay on a serial dilution of Eurasian and American H7 synthetic RNA after 60 minute incubation. NTC: no target control.

We designed and validated a SHINE assay specifically targeting Eurasian H7 AIV (SHINE-H7-Eurasian) (Figure 4B) and demonstrated its ability to discriminate against North American lineage at a range of tested concentrations (Figure 4C, Supplementary Figure 10). To assess broader specificity, we also tested this assay against a panel of unrelated vaccine-derived seasonal influenza viruses (Supplementary Figure 11) and observed no detectable cross-reactivity, further confirming its high specificity for H7 Eurasian strains. Additionally, to support deployment in field settings, we also adapted SHINE-H7-Eurasian to a lateral flow readout format, which maintained high lineage specificity (Figure 4D). Together, these results demonstrate the robustness of SHINE-H7-Eurasian, highlighting its potential for rapid, Eurasian lineage-specific H7 AIV detection in both laboratory and field settings.

## Discussion

Rising rates of AIV disease in wild animals, livestock, and humans underscore the growing risk of AIV-related health and economic impact, including significant pandemic potential should the capacity for human-to-human transmission evolve. The risk of an AIV pandemic can be mitigated if spillover events are detected early and public health action is taken, but limitations of available diagnostic assays have hindered the global response to widespread, increasing AIV disease activity. Here, we address unmet needs in AIV surveillance and detection by leveraging SHINE for AIV detection.

SHINE is highly specific and programmable, enabling existing avian influenza assays to be readily adapted to emerging viral strains and variants and diverse settings. Machine learning software platforms, such as ADAPT and BADGERS, greatly simplify and automate the design of new SHINE assays for different purposes as soon as viral genome sequences become publicly available (*37*, *43*). We developed highly sensitive and specific SHINE assays for the detection and surveillance of AIVs, including H5 subtype, clade 2.3.4.4b A(H5N1) and Eurasian H7 AIV. Compared to conventional RT-PCR, which remains the most widely used method for AIV diagnosis or surveillance, SHINE assays developed in this paper can be performed with minimal instrumentation under isothermal conditions at 37 ℃. SHINE assays developed in this study have a turnaround time between 60 and 120 minutes, and are compatible with lateral flow readouts, making them one step closer to point-of-need settings than conventional RT-PCR. In addition, SHINE-H5 achieved a LoD of 121.7 copies/μL on vaccine-derived H5 viral seedstocks, though not as sensitive as current PCR-based methods (*47*), significantly outperforming existing H5 immunoassay performance (*17*, *19*).

In addition to SHINE-H5, we also developed a clade-specific version (SHINE-H5-CS) targeting the 2.3.4.4b A(H5N1) lineage, which has dominated the global H5 phylogeny since 2021 and is associated with the ongoing cattle outbreak in the United States. SHINE H5-CS demonstrated strong sensitivity and high specificity across a wide range of input concentrations, with minimal off-target activity against non-2.3.4.4b H5. Given the ongoing spread of the clade 2.3.4.4b A(H5N1) strain across avian and mammalian hosts, SHINE-H5-CS provides the potential for lineage differentiation without the need for sequencing, enhancing the speed of clinical decision-making and public health response. Beyond clinical utility, such an assay also addresses a growing need for clade-level H5 typing tools in non-human surveillance contexts, where lineage resolution is increasingly prioritized for outbreak investigation and risk assessment.

We also designed and validated a Eurasian H7-specific SHINE assay (SHINE-H7-Eurasian) which reliably discriminated against North American H7 strains and unrelated seasonal influenza viruses. Like the SHINE-H5 assays, this assay is highly specific and compatible with lateral flow readout, making it possible for deployment in agricultural, wildlife, and environmental monitoring settings where discrimination between Eurasian and North American H7 lineages is critical.

While we demonstrated strong SHINE assay specificity using multiple vaccine-derived viral seedstocks and seasonal influenza-positive patient samples, access to H5 or Eurasian H7-positive clinical specimens remains limited, as confirmed human cases were rare. Future testing on such samples will be critical to further validate the clinical performance and real-world utility of the SHINE assays reported here. In addition, expanding assay compatibility to sample types from poultry, dairy products, or even environmental sources could broaden their utility for avian influenza surveillance in farms, wild bird markets, and rural clinics where laboratory infrastructure is limited.

## Methods

### Design of H5N1 and H7 Eurasian SHINE assays

H5N1 and H7 Eurasian sequences from 1980 to present day were compiled from both the GISAID EpiFlu and NCBI Influenza Virus databases and harmonized to ensure datasets were free from isolate duplicates. Alignment of these sequences was performed via MAFFT, using the ‘FFT-NS-1’ algorithm. RPA primers and crRNA guides were designed for SHINE-H5 and SHINE-H7-Eurasian using ADAPT (commit version #f202977), a machine learning based guide design software (*37*). The parameters were set as such: crRNA (oligo-length 28, mismatches 3, coverage-fraction 0.95, id-m 4) and RPA primer (oligo-length 30, mismatches 2, coverage-fraction 0.95, id-m 4), with maximize activity objective. The definition of these parameters are as follows: “oligo-length” refers to the length of the crRNA or RPA primer. “mismatches” mean the maximum number of tolerated mismatches between the oligonucleotide probes (oligos) and the target sequences. “coverage-fraction” sets minimum fraction of sequence diversity within the viral alignment that the oligos must capture according to the other user-defined criterion. “id-m” is one less than the minimum number of mismatches of the oligos with regards to an outgroup target in the cases of differential subtyping (a set of 5381 sequences were used as outgroup for H5 assay and 3801 for H7 Eurasian), ensuring that the oligos are type-specific. Designs were further evaluated through subsequent experiments.

### Design of 2.3.4.4b clade specific SHINE assays

8,737 clade 2.3.4.4.b A(H5N1) sequences and 498 non-2.3.4.4.b H5 sequences were compiled from the GISAID EpiFlu and NCBI Influenza Virus databases, spaning 2018 to present day. Sequences were aligned via MAFFT, using the ‘FFT-NS-1’ algorithm. These sequences were subsequently input into BADGERS to design crRNAs capable of specifically detecting clade 2.3.4.4b A(H5N1) sequences and discriminating against non-2.3.4.4b H5 sequences with single-nucleotide resolution. The “diff” objective (differentiation for variant identification) was selected, and both the “wgan-am” (i.e., Wasserstein generative adversarial network–activation maximization) and “evolutionary” algorithms were employed for crRNA design. These algorithms search over a landscape of guide sequences to generate maximally-fit guides. Candidates with the highest predicted activity and compatible with the RPA primers from SHINE-H5 were selected for further evaluation and testing.

### Nucleic acids, materials, and reagents

All RPA primers were ordered as custom DNA oligos (IDT). crRNAs were ordered as synthetic Alt-R CRISPR RNAs (IDT). gBlock DNA gene fragments (IDT) with a T7 promoter were ordered as synthetic testing targets. gBlocks were *in vitro* transcribed (IVT) using HiScribe™ T7 High Yield RNA Synthesis Kits (NEB) to create synthetic RNA targets. A 30uL IVT master mix was made consisting of 1x Reaction Buffer (NEB), 10mM ATP, 10mM UTP, 10mM CTP, 10mM GTP, 1ug gBlock DNA. IVT mastermix is incubated overnight at 37℃.

Following transcription, 2uL DNAse I (NEB) and 18ul H2O were added to each reaction to digest the DNA template. Mastermix was incubated at 37℃ for 30 minutes and reaction was terminated by adding 5 uL of 0.5M EDTA. Synthetic RNA was purified with RNACLEAN XP (Beckman) according to the manufacturer’s instructions quantified with Nanodrop. All nucleic acids were diluted to the desired concentration with nuclease-free water. See Supplementary Table 4, 5 for sequence details.

### Viral seedstock extraction and quantification

All vaccine-derived virus seedstocks were provided by CDC, see Supplementary Table 3 for sequence detail. Viral seedstocks were extracted with MagMAX™ CORE Nucleic Acid Purification Kit (Thermo) with KingFisher Flex non-heated script according to manufacturer instructions, using a starting volume of 200 μl per sample and eluted into 90 μl of nuclease-free water. RT-qPCR reactions were prepared with TaqPath™ 1-Step RT-qPCR Master Mix, CG (Thermo) according to manufacturer instructions, using a primer concentration of 500nM and a probe concentration of 200nM. Reaction mixes were incubated on a QuantStudio 6 (Applied Biosystems) with the following cycling conditions: hold at 25°C for 2min, reverse transcription at 50 °C for 15 min, polymerase activation at 95 °C for 2 min and 40 cycles with a denaturing step at 95 °C for 3 s followed by annealing and elongation steps at 60 °C for 30 s. Data were analyzed using the Standard Curve module of the Applied Biosystems analysis software. Quantified viral seedstocks were spiked into universal transport medium (BD) to desired concentration for the LoD and Specificity experiments.

### Prototype AIV SHINE assay

This protocol involved the amplification and detection of nucleic acids in a single reaction. A SHINE master mix was made consisting of 1X SHINE buffer (20 mM HEPES pH 8.0 with 60 mM KCl and 5% PEG-8000), 45 nM LwaCas13a protein, 62.5 nM 6rU FAM quenched reporter, 1 unit/µL murine RNase inhibitor, 2 mM of each rNTP, 1 unit/µL NextGen T7 RNA polymerase, and 120 nM forward and reverse RPA primers. This master mix was used to resuspend the TwistAmp Basic RPA pellets (1 pellet per 102 µL master mix). After RPA pellet resuspension, 2 units/µL SuperScript IV reverse transcriptase, 0.1 unit/µL RNase H (NEB) and 45 nM crRNA were added to the mastermix. Finally, magnesium acetate was added to reach a concentration of 14 mM to create the final master mix.

Samples were added in a 1:9 ratio with the final master mix for a final volume of 20µL. For the fluorescence-based readout, the fluorescence of each reaction was measured using a Cytation 5 plate reader (Agilent) with a monochromator, set to excitation at 485 nm and emission at 520 nm with a 20 nm monochromator width. Readings were recorded every 5 minutes at 37°C for 90 minutes.

### H5 SHINE assay optimization

Optimization experiments were performed by adding a non-T7 attached forward primer at different ratio and varying individual components of the prototype H5 SHINE assay while keeping all other conditions constant to the prototype version. The following ratio for T7-attached primer to non-T7attached primer was tested at 1:2.5, 1:3, 1:3.5, 1:4. Overall primer concentration was tested at 140, 160 and 180nM. RNase H concentration were tested at 0, 0.1 and 1 units/μL. FAM quenched reporter concentration was tested at 62.5, 78.2, 93.7nM, magnesium acetate concentration was tested at 14, 18, 22mM. Fluorescence signals at 60 minutes were used as the primary criterion for selecting optimal reagent concentrations. A final assay formulation was prepared by combining the individually optimized concentrations of all reagents.

### Optimized H5 SHINE assay and lateral flow readout adaptation

A SHINE master mix was made consisting of 1X SHINE buffer (20 mM HEPES pH 8.0 with 60 mM KCl and 5% PEG-8000), 45 nM LwaCas13a protein, 78.2 nM 6rU FAM quenched reporter (for fluorescence readout) or 1µM 14rU FAM-Biotin reporter (for paper-based readout), 1 unit/µL murine RNase inhibitor, 2 mM of each rNTP, 1 unit/µL NextGen T7 RNA polymerase, 40 nM T7-attached forward RPA primer, 120 nM non-T7-attached forward RPA primer, and 160nM reverse RPA primer. This master mix was used to resuspend the TwistAmp Basic RPA pellets (1 pellet per 102 µL master mix). After RPA pellet resuspension, 2 units/µL SuperScript IV reverse transcriptase, 0.1 unit/µL RNase H (NEB) and 45 nM crRNA were added to the mastermix. Finally, magnesium acetate was added to reach a concentration of 14 mM to create the final master mix.

Samples were added in a 1:9 ratio with the final master mix for a final volume of 20µL (for fluorescence readout) or 40µL (for paper-based readout). For fluorescent readout, reads are taken similar to the prototype assay for 60 minutes. For the paper-based readout, the reactions were incubated at 37°C for 60 minutes, and then diluted 1:4 in HybriDetect Assay Buffer (Milenia Biotec). The buffered solution was incubated at room temperature for 5 minutes; after which, a HybriDetect1 lateral flow paper strip was added. Test images were taken 2 minutes after the strip was added, using a smartphone (iPhone 13 pro) camera. Lateral flow reactions that are positive for the template show a top test (T) fluorescence band on the paper strip after incubation, and they may or may not show a bottom control (C) band (*34*, *36*). Lateral flow reactions that are negative for the template only show a bottom (C) band. Lateral flow reactions are not designed to be quantitative.

### Clinical samples ethics statement and characterization

Clinical specimens were obtained from the Massachusetts General Hospital (MGH) Clinical Microbiology Laboratory between January 12 and March 13, 2025. All samples utilized in this study were convenience specimens collected during routine clinical care and were considered excess materials that would otherwise have been discarded. Ethical review and approval for the use of these specimens was provided by the Massachusetts General Brigham Institutional Review Board (MGB IRB Protocol #2019P003305).

All specimens consisted of nasopharyngeal (NP) swabs eluted in viral transport medium that had previously tested positive for influenza A or B by standard clinical nucleic acid amplification testing (NAAT). Depending on the clinical assay used, the results included either a quantitative cycle threshold (Ct) value from quantitative PCR (qPCR; Xpert® Xpress CoV-2/Flu/RSV plus, Cepheid, Sunnyvale, CA) or identification of influenza A subtypes (H1-2009 or H3; BIOFIRE® Respiratory 2.1 (RP2.1) Panel, bioMérieux, Salt Lake City, UT). Nucleic acid was extracted from 200 µL of NP swab eluate using the MagMax Prime Viral/Pathogen Nucleic Acid Isolation Kit on the KingFisher Flex platform (Thermo Fisher Scientific, Waltham, MA, USA), following the manufacturer’s recommended protocol. Extracted specimens were subsequently confirmed positive for influenza A or B utilizing the CDC’s Influenza SARS-CoV-2 Multiplex Assay performed on a QuantStudio 6 Pro Real-Time PCR instrument (Thermo Fisher Scientific, Waltham, MA, USA). Reactions were run in triplicate in a final volume of 20µL per replicate on the QuantStudio™ 6 Pro (ThermoFisher Scientific, Waltham, MA). Each reaction contained 5µL of 4X TaqPath™ 1-Step RT-qPCR Master Mix, CG (ThermoFisher Scientific, Waltham, MA) 5µL of template nucleic acid, 800nM final concentration of each primer, and 200nM final concentration of probe (*48*). Positive influenza A specimens were further screened for H5 using a previously published qPCR assay (*49*).

Characterized NP swabs were extracted using the MagMax Prime Viral/Pathogen Nucleic Acid Isolation Kit on the KingFisher Flex platform (Thermo Fisher Scientific, Waltham, MA, USA), following the manufacturer’s recommended protocol. This new extraction was then tested using SHINE-H5 or SHINE-H5-LFA for comparison.

### Data analysis

Plots and graphs were generated using Prism software unless otherwise specified.

### Sequence conservation

Aligned sequences of H5N1 in FASTA format were parsed using the Biopython library (v1.84) and similarity was assessed at each nucleotide position across all sequences in the alignment. For each position, the most frequent nucleotide was identified as consensus nucleotide and its frequency across sequences was used to compute the percentage similarity at the position. The percentage conservation was defined as the proportion of sequences matching the consensus base at that position, normalized to the total number of sequences.

### Detection and SHINE assay with fluorescent readout

Fluorescence values are the mean ± standard deviation of 3 technical replicates. For clinical specificity experiments (Figure 2C), fluorescence values are normalized to the mean fluorescence of extracted H5N1 seedstock at 10^2^ copies/μL in each sample batch.

### Limit of detection

Detection probability data were analyzed and plotted in Python using the statsmodels (v0.14.0) and matplotlib (v3.7.1) packages. A simple logistic regression model was fitted using the Generalized Linear Model function in statsmodels to estimate the probability of detection as a function of H5N1 viral seedstock concentration. The log-transformed concentrations were used as the independent variable and the number of positive and negative test results at each concentration were modeled as a binomial outcome. The 95% limit of detection (LoD95) was defined as the predicted concentration for 95% probability of detection. Wald 95% confidence intervals were computed based on the standard error of the fitted model and visualized as a shaded region around the logistic curve to indicate the uncertainty of the estimated detection probabilities.

### Phylogeny & sequence frequency H5 Clade Frequency

H5 sequences from North America and Eurasia were collected from GISAID between 2015 to present day. GISAID annotations were used to calculate the five most common H5 clades represented in these sequences. The relative frequency for each clade was subsequently calculated using the Biopython library (v.184) and plotted per year.

### H7 Phylogeny

H7 sequences from North America and Eurasia were collected from GISAID and clustered using MeShClust v3.0 at the 0.85 identity score threshold. The most representative sequence was extracted from each cluster, aligned via MAFFT (‘FFT-NS-i’ algorithm), and input into FastTree (v.2.2.0) for phylogenetic analysis. Phylogenetic relationships between sequences were visualized in Python using the ETE v3 (v4.3.0) package.

## Data Availability

All data produced in the present study are available upon reasonable request to the authors.

## Acknowledgements

We would like to thank members of the Myhrvold laboratory, Lemieux laboratory, Sabeti laboratory and the CDC’s Influenza Genomics lab - notably, J. Devlin for handling patient sample extractions, S. Garg for helping perform assays on SHINE-H5 specificity validation, S. Mantena for his contributions to initial H5 clade specific crRNA design; J. Barnes and M. Kirby for kindly providing viral seedstocks used in this study; and those researchers and laboratories who have made influenza sequencing data publicly available, which informed our assay design.

We acknowledge funding from the Centers for Disease Control and Prevention award 75D30122C15113 NIH R01 AI182281, and from the Princeton Catalysis Initiative (all to C.M.), Clinical sample collection and management was supported by CDC NU50CK000629 (Pathogen Genomic Center of Excellence, to J.E.L.)

## Author contributions

C.M. conceived the study. A.G. and Y.H. designed assays in silico. Y.H. performed experiments and data analysis to develop and validate SHINE-H5, SHINE-H5-CS, and SHINE-H7-Eurasian assays. A.G. constructed phylogenies for H5N1 and H7. J.E.L. and G.A. provided critical insights on clinically relevant experiments. Y.H. and A.G. wrote the paper with guidance from C.M. All authors reviewed the manuscript.

## Conflict of interest/competing interests

C.M. is listed as an inventor on patent applications related to the SHINE technology. Y.H., A.G., C.M. are listed as co-inventors on a patent application relating to this study. No other disclosures are reported.

