## Supplementary Information for "Streamlined CRISPR-based assays for detection and subtyping of avian influenza"

**Supplementary Table 1**. Summary of current CRISPR-Dx for AIV detection.

| Reference | Combined (Y/N) | Target gene | Input | Subtyping/clade typing | Cas protein(s) | Sample-to-answer time (min) | Analytical sensitivity (cp/µL) | | Off-target seedstocks tested | Clinical Sensitivity (%) | Clinical Specificity (%) |
|---|---|---|---|---|---|---|---|---|---|---|---|
| | | | | | | | Fluorescence | LFA | | | |
| (*27*) | N | HA | RNA | H5 | LwaCas13a | 50 | N.D. | 1 | H3, H7, H9, H10-AIV, IBV, NDV, RVA and DAstV | 81.6 | 100 |
| (*28*) | N | M | cDNA | No | LwaCas13a | N.D. | 69 | 690 | NDV, IBV, IBDV | N.D. | N.D. |
| (*29*) | N | NP | cDNA/ plasmid | No | LwaCas13a | 60 | N.D. | 1 | NDV, IBV, ILTV | N.D. *** | N.D. |
| (*30*) | N | M, NP | RNA | No | LbaCas12a | 150 | 6.7 (M) ,12 (NP) | 67 - (M) , $1.2\times10^4$ (NP) | NDV, IBV, IBDV** | 98.1 | 96 |
| (*31*) | N | HA | RNA | H9N2 | LwaCas13a | 60-90* | 1 | N.D. | H5N1 AIV, H1N1 AIV, NDV, IBDV, and IBV | N.D. | N.D. |
| (*32*) | N | HA | RNA | H5 | LbaCas12a | 60-180 | 1.9 | $1.9\times10^3$ | 16 subtypes of AIVs (H1-H16), NDV, IBV, IBDV | 93.75 | 84.84 |
| (*33*) | N | HA | RNA | H5 | LbaCas12a | 60 | 100 $EID_{50}$/mL | 1000 $EID_{50}$/mL | H9N2, H1N1, H3N2, IBV, H1N5, H7N5, H6N2, H11N2, H10N7 | 80 | 100 |
| This study | Y | HA | RNA | H5, 2.3.4.4b A (H5N1), H7 Eurasian | LwaCas13a | 60 | 121.7 | 25 | H3, H7N9, pdmH1N1, VIC, YAM | N.D. | 100**** |

* estimated through kinetics
** compared to in vitro transcribed M/NP gene of these viruses
*** 98.75% coincidence rate compared to agarose gel after PCR
**** compared to human samples infected with seasonal IAV
Acronyms: Newcastle disease virus (NDV), Infectious bronchitis virus (IBV), Infectious bursal disease virus (IBDV)
N.D.: not determined

**Supplementary Table 2**. Strain information for cross-reactivity panel for Figure 3E, Supplementary Figure 5A, 11.

| Sequence name | HA segment origin |
|---|---|
| pdmH1N1 | A/Michigan/45/2015 |
| A/H3 | A/Singapore/INFIMH-16-0019/2016 |
| A/H3 Variant | A/Ohio/28/2016 |
| A/H5N1 | A/gyrfalcon/Washington/41088-6/2014 |
| A/H7N9 | A/HongKong/125/2017-IDCDC-RG33A |
| B/VIC | B/Colorado/06/2017 |
| B/YAM | B/Phuket/3073/2013 |

**Supplementary Table 3**. Patient sample information for Figure 2C, Supplementary Figure 5B, 5C, 7.

| Sample ID | Target 1 | Target 2 | Target 3 | CT FLU B | CT FLU A1 | CT FLU A2 | Average SHINE fluorescence (a.u.) | Normalized fluorescence |
|---|---|---|---|---|---|---|---|---|
| 1 - positive control* | N/A | N/A | N/A | N/A | N/A | N/A | 2529.7 | 1.000 |
| 1 - 1 | INFLUENZA A | INFLUENZA B | 0 | 24.5 | 20.2 | 23.7 | 579.0 | 0.229 |
| 1 - 2* | INFLUENZA A | INFLUENZA A H1-2009 | INFLUENZA A H3 | 0 | 0 | 0 | 596.7 | 0.236 |
| 1 - 3 | INFLUENZA A | INFLUENZA A H3 | 0 | 0 | 0 | 0 | 574.3 | 0.227 |
| 1 - 4 | INFLUENZA A | INFLUENZA A H3 | 0 | 0 | 0 | 0 | 602.0 | 0.238 |
| 1 - 5 | INFLUENZA B | 0 | 0 | 20.1 | 0 | 0 | 628.3 | 0.248 |
| 1 - 6 | INFLUENZA A | INFLUENZA A H3 | 0 | 0 | 0 | 0 | 615.0 | 0.243 |
| 1 - 7* | INFLUENZA A | INFLUENZA A H3 | 0 | 0 | 0 | 0 | 562.3 | 0.222 |
| 1 - 8* | INFLUENZA A | 0 | 0 | 0 | 25.8 | 27.3 | 587.7 | 0.232 |
| 1 - 9 | INFLUENZA A | 0 | 0 | 0 | 31.2 | 34.6 | 604.7 | 0.239 |
| 1 - 10 | INFLUENZA A | 0 | 0 | 0 | 22 | 23.4 | 458.7 | 0.181 |
| 1 - 11 | INFLUENZA A | INFLUENZA A H3 | 0 | 0 | 0 | 0 | 632.0 | 0.250 |
| 1 - 12* | INFLUENZA B | 0 | 0 | 28 | 0 | 0 | 548.7 | 0.217 |
| 1 - 13 | INFLUENZA A | 0 | 0 | 0 | 21.1 | 21.8 | 629.7 | 0.249 |
| 1 - 14* | INFLUENZA A | INFLUENZA A H1-2009 | INFLUENZA A H3 | 0 | 0 | 0 | 580.0 | 0.229 |
| 1 - 15* | INFLUENZA A | INFLUENZA B | 0 | 11.8 | 34.6 | 0 | 532.7 | 0.211 |
| 1 - 16 | INFLUENZA B | 0 | 0 | 21.1 | 0 | 0 | 643.0 | 0.254 |
| 1 - 17 | INFLUENZA A | 0 | 0 | 0 | 31.1 | 31.9 | 614.3 | 0.243 |
| 1 - 18 | INFLUENZA A | 0 | 0 | 0 | 25.4 | 26.6 | 542.3 | 0.214 |
| 1 - 19 | INFLUENZA A | 0 | 0 | 0 | 27.4 | 28.7 | 637.3 | 0.252 |
| 1 - 20 | INFLUENZA B | 0 | 0 | 38.8 | 0 | 0 | 636.3 | 0.252 |
| 1 - 21 | INFLUENZA B | 0 | 0 | 32.2 | 0 | 0 | 637.3 | 0.252 |
| 1 - 22 | INFLUENZA A | 0 | 0 | 0 | 22.4 | 24.1 | 627.3 | 0.248 |

| | | | | | | | | |
|---|---|---|---|---|---|---|---|---|
| 1 - 23 | INFLUENZA A | INFLUENZA A H3 | 0 | 0 | 0 | 0 | 695.7 | 0.275 |
| 1 - 24 | INFLUENZA A | INFLUENZA A H3 | 0 | 0 | 0 | 0 | 604.7 | 0.239 |
| 1 - 25* | INFLUENZA A | INFLUENZA A H3 | 0 | 0 | 0 | 0 | 581.3 | 0.230 |
| 1 - 26 | INFLUENZA A | 0 | 0 | 0 | 20.8 | 22.1 | 424.7 | 0.168 |
| 1 - 27 | INFLUENZA A | 0 | 0 | 0 | 21.6 | 23.3 | 569.7 | 0.225 |
| 1 - 28 | INFLUENZA A | INFLUENZA A H3 | 0 | 0 | 0 | 0 | 647.7 | 0.256 |
| 1 - 29 | INFLUENZA B | 0 | 0 | 35.1 | 0 | 0 | 643.3 | 0.254 |
| 1 - 30 | INFLUENZA B | 0 | 0 | 33.7 | 0 | 0 | 629.3 | 0.249 |
| 1 - 31 | INFLUENZA A | 0 | 0 | 0 | 25.7 | 27.1 | 640.7 | 0.253 |
| 1 - 32 | INFLUENZA A | 0 | 0 | 0 | 34.8 | 34.7 | 642.3 | 0.254 |
| 1 - NTC* | N/A | N/A | N/A | N/A | N/A | N/A | 631.0 | 0.249 |
| 2 - positive control | N/A | N/A | N/A | N/A | N/A | N/A | 1262.3 | 1.000 |
| 2 - 1* | INFLUENZA A | 0 | 0 | 0 | 33.5 | 33.2 | 383.3 | 0.304 |
| 2 - 2 | INFLUENZA B | 0 | 0 | 33.2 | 0 | 0 | 392.7 | 0.311 |
| 2 - 3 | INFLUENZA B | 0 | 0 | 30.5 | 0 | 0 | 382.0 | 0.303 |
| 2 - 4 | INFLUENZA A | 0 | 0 | 0 | 36.9 | 0 | 380.0 | 0.301 |
| 2 - 5 | INFLUENZA A | 0 | 0 | 0 | 36.3 | 37.1 | 400.3 | 0.317 |
| 2 - 6 | INFLUENZA A | INFLUENZA A H1-2009 | 0 | 0 | 0 | 0 | 398.0 | 0.315 |
| 2 - 7 | INFLUENZA A | INFLUENZA A H1-2009 | 0 | 0 | 0 | 0 | 385.7 | 0.306 |
| 2 - 8 | INFLUENZA A | INFLUENZA A H1-2009 | 0 | 0 | 0 | 0 | 384.7 | 0.305 |
| 2 - 9 | INFLUENZA A | INFLUENZA A H1-2009 | 0 | 0 | 0 | 0 | 407.0 | 0.322 |
| 2 - 10 | INFLUENZA A | INFLUENZA A H1-2009 | 0 | 0 | 0 | 0 | 401.0 | 0.318 |
| 2 - 11 | INFLUENZA B | 0 | 0 | 29.4 | 0 | 0 | 408.7 | 0.324 |
| 2 - 12 | INFLUENZA A | INFLUENZA A H1-2009 | 0 | 0 | 0 | 0 | 377.3 | 0.299 |
| 2 - 13* | INFLUENZA A | INFLUENZA A H1-2009 | 0 | 0 | 0 | 0 | 417.0 | 0.330 |

| | | | | | | | | |
|---|---|---|---|---|---|---|---|---|
| 2 - 14 | INFLUENZA A | INFLUENZA A H1-2009 | 0 | 0 | 0 | 0 | 416.0 | 0.330 |
| 2 - 15 | INFLUENZA A | 0 | 0 | 0 | 35.9 | 37.2 | 417.3 | 0.331 |
| 2 - 16 | INFLUENZA A | 0 | 0 | 0 | 36 | 0 | 404.7 | 0.321 |
| 2 - 17 | INFLUENZA B | 0 | 0 | 24.8 | 0 | 0 | 340.7 | 0.270 |
| 2 - 18 | INFLUENZA A | 0 | 0 | 0 | 36.6 | 38 | 406.3 | 0.322 |
| 2 - 19 | INFLUENZA B | 0 | 0 | 36.7 | 0 | 0 | 422.3 | 0.335 |
| 2 - 20 | INFLUENZA A | 0 | 0 | 0 | 31.4 | 33.9 | 413.7 | 0.328 |
| 2 - 21 | INFLUENZA B | 0 | 0 | 38.6 | 0 | 0 | 424.3 | 0.336 |
| 2 - 22 | INFLUENZA B | 0 | 0 | 35.3 | 0 | 0 | 401.7 | 0.318 |
| 2 - 23 | INFLUENZA B | 0 | 0 | 26.3 | 0 | 0 | 409.0 | 0.324 |
| 2 - 24 | INFLUENZA B | 0 | 0 | 24.3 | 0 | 0 | 410.3 | 0.325 |
| 2 - 25 | INFLUENZA B | 0 | 0 | 31.5 | 0 | 0 | 446.7 | 0.354 |
| 2 - 26* | INFLUENZA B | 0 | 0 | 26 | 0 | 0 | 394.3 | 0.312 |
| 2 - 27* | INFLUENZA A | INFLUENZA A H3 | INFLUENZA B | 0 | 0 | 0 | 416.3 | 0.330 |
| 2 - 28 | INFLUENZA B | 0 | 0 | 23.4 | 0 | 0 | 431.7 | 0.342 |
| 2 - 29 | INFLUENZA B | 0 | 0 | 25.8 | 0 | 0 | 404.0 | 0.320 |
| 2 - 30* | INFLUENZA A | INFLUENZA B | 0 | 18.2 | 30.3 | 30.4 | 382.3 | 0.303 |
| 2 - 31 | INFLUENZA A | 0 | 0 | 0 | 27.6 | 28.4 | 421.0 | 0.334 |
| 2 - 32* | INFLUENZA A | INFLUENZA A H1-2009 | 0 | 0 | 0 | 0 | 420.7 | 0.333 |
| 2 - NTC | N/A | N/A | N/A | N/A | N/A | N/A | 440.7 | 0.349 |

* Samples chosen for SHINE-H5-LFA specificity validation

**Supplementary Table 4**. Primer, crRNA, and reporter sequences ordered through IDT.

| Name | Oligo type | Sequence | Assays used |
|---|---|---|---|
| H5_Design1_RPA_T7_F | custom DNA oligo | GAAATTAATACGACTCACTATAGGGGAGAGTAATGGAAATTTCATTGCTCCAGAA | Prototype SHINE H5, SHINE-H5, SHINE-H5-LFA |
| H5_Design1_RPA_R | custom DNA oligo | GGAATGGCATACTAGAGTTTATCGCCCCTA | Prototype SHINE H5, SHINE-H5, SHINE-H5-LFA |
| H5_Design1_RPA_nonT7_F | custom DNA oligo | GAGAGTAATGGAAATTTCATTGCTCCAGAA | SHINE-H5, SHINE-H5-LFA |
| H5_CladeSpecific_RPA_F | custom DNA oligo | GAAATTAATACGACTCACTATAGGGTTTCGAGAGTAATGGAAATTTCATTGCTCCA | 2.3.4.4b clade-specific H5 SHINE |
| H5_CladeSpecific_RPA_R | custom DNA oligo | AATGGCATACTAGAATTTATCGCTCCTA | 2.3.4.4b clade-specific H5 SHINE |
| H7_Eurasian_RPA_T7_F | custom DNA oligo | GAAATTAATACGACTCACTATAGGGTCAAACTAAGCAGCGGCTACAAAGATGTGA | SHINE-H7-Eurasian |
| H7_Eurasian_RPA_T7_R | custom DNA oligo | AGGCCCATTACAATGGCTAGAAGTATGAAA | SHINE-H7-Eurasian |
| H7_Eurasian_RPA_nonT7_F | custom DNA oligo | TCAAACTAAGCAGCGGCTACAAAGATGTGA | SHINE-H7-Eurasian |
| FAM-6U-Q reporter | custom RNA oligo | /56-FAM/rUrUrUrUrUrU/3IABkFQ/ | Prototype SHINE H5, SHINE-H5, SHINE-H7-Eurasian |
| FAM-14U-Bio reporter | custom RNA oligo | /5BiosG/rUrUrUrUrUrUrUrUrUrUrUrUrUrU/36-FAM/ | SHINE-H5-LFA, SHINE-H7-Eurasian-LFA |
| H5_Design1_crRNA | CRISPR Custom Guide RNA | /AltR1/rGrArUrUrUrArGrArCrUrArCrCrCrCrArArArArArCrGrArArGrGrGrGrArCrUrArArArArCrCrCrCrCrUrUrUrCrUrUrGrArCrArArUrUrUrUrGrUrArUrGrCrArUrA/AltR2/ | Prototype SHINE H5, SHINE-H5, SHINE-H5-LFA |
| H5_Design2_crRNA | CRISPR Custom Guide RNA | /AltR1/rGrArUrUrUrArGrArCrUrArCrCrCrCrArArArArArCrGrArArGrGrGrGrArCrUrArArArArCrGrCrArGrUrArUrUrCrArGrArArGrArArGrCrArArGrArUrUrArArArA/AltR2/ | Prototype SHINE H5 |
| H5_CladeSpecific_Design1_crRNA | CRISPR Custom Guide RNA | /AltR1/rGrArUrUrUrArGrArCrUrArCrCrCrCrArArArArArCrGrArArGrGrGrGrArCrUrArArArArCrUrUrUrGrGrUrGrUrUrGrCrArGrUrGrGrCrCrArUrArUrUrCrCrArCrU/AltR2/ | Prototype 2.3.4.4b clade-specific H5 SHINE |
| H5_CladeSpecific_Design2_crRNA | CRISPR Custom Guide RNA | /AltR1/rGrArUrUrUrArGrArCrUrArCrCrCrCrArArArArArCrGrArArGrGrGrGrArCrUrArArArArCrUrUrGrArCrArUrUrUrGrGrUrGrUrUrGrCrArGrUrGrGrCrCr | Prototype 2.3.4.4b clade-specific H5 SHINE |

| | | | |
|---|---|---|---|
| | | ArUrArU/AltR2/ | |
| H5_CladeSpecific_Design3_crRNA | CRISPR Custom Guide RNA | /AltR1/rGrArUrUrUrArGrArCrUrArCrCrCrCrArArArArArCrGrArArGrGrGrArCrUrArArArArCrUrUrGrGrUrGrUrUrGrCrArGrUrGrGrCrCrArUrArUrUrCrCrArCrUrC/AltR2/ | Prototype 2.3.4.4b clade-specific H5 SHINE, SHINE-H5-CS |
| H7_Eurasian_crRNA | CRISPR Custom Guide RNA | /AltR1/rGrArUrUrUrArGrArCrUrArCrCrCrCrArArArArArCrGrArArGrGrGrArCrUrArArArArCrCrArUrGrArUrGrCrCrCrCrGrArArGrCrUrArArArCrCrArArArGrUrA/AltR2/ | SHINE-H7-Eurasian |

**Supplementary Table 5**. gBlock sequences ordered through IDT.

| Name | Oligo type | Sequence | Assays used | Strain origin |
|---|---|---|---|---|
| H5_strain1_full | gBlocks Gene Fragment | GAAATTAATACGACTCACTATAGGGATGGAGAACATAGTACTTCTTCTTGCAATAGTTAGCCTTGTTAAAAGTGATCAGATTTGCATTGGTTACCATGCAAACAATTCGACAGAGCAAGTTGACACGATAATGGAAAAGAACGTCACTGTTACACATGCCCAAGACATACTGGAAAAAACACACAACGGGAAGCTCTGTGATCTAAATGGGGTGAAGCCTCTGATTTTAAAGGATTGTAGTGTAGCTGGATGGCTCCTCGGAAACCCAATGTGCGACGAATTCATCAGAGTGCCGGAATGGTCCTACATAGTGGAGCGGACTAATCCAGCTAATGACCTCTGTTACCCAGGGAGCCTCAATGACTATGAAGAACTGAAACACCTGTTGAGCAGAATAAATCATTTTGAGAAGATTCTGATCATCCCCAAGAGTTCCTGGCCAAATCATGAAACATCACTAGGGGTGAGCGCAGCTTGTCCATACCAGGGAGCGCCCTCCTTTTTCAGAAATGTGGTGTGGCTTATCAAAAAGAACGATGCATACCCAACAATAAAGATAAGCTACAATAATACCAATCGGGAAGATCTCTTGATACTGTGGGGGATTCATCATTCCAACAATGCAGAAGAGCAGACAAATCTCTACAAAAACCCAACCACCTACATTTCAGTTGGAACATCAACTTTAAACCAGAGGTTGGTACCAAAAATAGCTACTAGATCCCAAGTAAACGGGCAACGTGGAAGAATGGACTTCTTCTGGACAATTTTAAAACCAGATGATGCAATCCATTTCGAGAGTAATGGAAATTTCATTGCTCCAGAATATGCATACAAAATTGTCAAGAAAGGGGACTCAACAATTATGAAAAGTGGAGTGGAATATGGCCACTGCAACACCAAATGTCAAACCCCAGTAGGAGCGATAAATTCTAGTATGCCATTCCACAACATACATCCTCTCACCATTGGGGAATGCCCCAAATATGTGAAGTCAAACAAGTTGGTCCTTGCGACTGGGCTCAGAAATAGTCCTCTAAGAGAAAAGAGAAGAAAAAGAGGCCTGTTTGGGGCGATAGCAGGGTTTATAGAGGGAGGATGGCAGGGAATGGTTGATGGTTGGTATGGGTACCATCATAGCAATGAGCAGGGGAGTGGGTACGCTGCAGACAAAGAATCCACCCAAAAGGCAATAGATGGAGTTACCAATAAGGTCAACTCAATCATTGACAAAATGAACACTCAATTTGAGGCAGTTGGAAGGGAGTTTAATAACTTAGAAAGGAGGATAGAGAATTTGAACAAGAAAATGGAAGACGGATTCCTAGATGTCTGGACCTATAATGCTGAACTTCTAGTTCTCATGGAAAACGAGAGGACTCTAGATTTCCATGATTCAAATGTCAAGAACCTTTACGACAAAGTCAGACTACAGCTTAGGGATAATGCAAAGGAGCTGGGTAACGGCTGTTTCGAATTCTATCACAAATGCGATAATGAATGTATGGAAAGTGTGAGAAATGGGACGTATGACTACCCTCAGTATTCAGAAGAAGCAAGATTAAAAAGAGAAGAAATAAGCGGAGTGAAATTAGAATCAATAGGAACTTACCAGATACTGTCAATTTATTCAACAGCGGCGAGTTCCCTAGCACTGGCAATCATGATGGCTGGTCTATCTTTATGGATGTGCTCCAATGGGTCGTTACAGTGCAGAATTTGCATCTAG | Prototype SHINE H5 | \|A/white-tailed_eagle/Estonia/2021\|2021-05-16\|1945381 |

<table>
<tr><td>H5_strain2_full</td><td>gBlocks Gene Fragment</td><td>GAAATTAATACGACTCACTATAGGGATGGAGAACATAGTAC<br>TTCTTCTTGCAATAGTTAGCCTTGTTAAAAGTGATCAGATT<br>TGCATTGGTTACCATGCAAACAATTCGACAGAACAGGTTGA<br>CACGATAATGGAAAAGAACGTCACTGTTACACATGCCCAAG<br>ACATACTGGAAAAAACACACAACGGGAAGCTCTGTGATTTA<br>AATGGGGTGAAGCCTCTGATTTTAAAGGATTGTAGTGTAGC<br>TGGATGGCTCCTCGGAAACCCAATGTGCGACGAATTCATCA<br>GAGTGCCGGAATGGTCCTACATAGTGGAGCGGGCTAATCCA<br>GCTAATGACCTCTGTTACCCAGGGAGCCTCAATGACTATGA<br>AGAACTGAAACACCTGTTGAGCAGAATAAATCATTTTGAGA<br>AGATTCTTATCATCCCCAAGAGTTCTTGGCCAAATCATGAA<br>ACATCACTAGGGGTGAGCGCAGCTTGTCCATACCAGGGAGC<br>GCCCTCCTTTTTCAGAAATGTGGTGTGGCTTATCAAAAAGA<br>ACGATGCATACCCAACAATAAAGATAAGCTACAATAATACC<br>AATAGGGAAGATCTCTTGATACTGTGGGGGATTCATCATTC<br>CAACAATGCAGAAGAACAGACAAATCTCTATAAAAACCCAA<br>CCACCTACATTTCAGTTGGAACATCAACTTTAAACCAGAGG<br>TTGGTACCAAAAATAGCTACTAGATCCCAAGTAAACGGGCA<br>ACGTGGAAGAATGGACTTCTTCTGGACAATTTTAAAACCAG<br>ATGATGCAATCCATTTCGAGAGTAATGGAAATTTCATTGCT<br>CCAGAATATGCATATAAAATTGTCAAGAAAGGGGACTCAAC<br>AATTATGAAAAGTGGAGTGGAATATGGCCACTGCAACACCA<br>AATGTCAAACCCCAGTAGGAGCGATAAATTCTAGTATGCCA<br>TTCCACAACATACATCCTCTCACCATTGGGGAATGCCCCAA<br>ATACGTGAAGTCAAACAAGTTGGTCCTTGCGACTGGGCTCA<br>GAAATAGTCCTCTAAGAGAAAAGAGAAGAAAAAGAGGCCTG<br>TTTGGGGCGATAGCAGGGTTTATAGAGGGAGGATGGCAGGG<br>AATGGTTGATGGTTGGTATGGGTACCATCATAGCAATGAGC<br>AGGGGAGTGGGTATGCTGCAGACAAAGAATCTACCCAAAAG<br>GCAATAGATGGAGTTACCAATAAGGTCAACTCAATCATTGA<br>CAAAATGAACACTCAATTTGAGGCAGTTGGAAGGGAGTTTA<br>ATAACTTAGAAAGGAGGATAGAGAATTTGAACAAGAAAATG<br>GAAGACGGATTCCTAGATGTCTGGACCTATAATGCTGAACT<br>TCTAGTTCTCATGGAAAACGAGAGGACTCTAGATTTCCATG<br>ATTCAAATGTCAAGAACCTTTACGACAAAGTCAGACTACAG<br>CTTAGGGATAATGCAAAGGAGCTGGGTAATGGCTGTTTCGA<br>ATTCTATCACAAATGCGATAATGAATGTATGGAAAGTGTGA<br>GAAATGGGACGTATGACTACCCTCAGTATTCAGAAGAAGCA<br>AGGTTAAAAAGAGAAGAAATAAGCGGAGTGAAATTAGAATC<br>AATAGGAACTTACCAGATACTGTCAATTTATTCAACAGCGG<br>CGAGTTCCCTAGCACTGGCAATCATGATAGCTGGTCTATCT<br>TTATGGATGTGCTCCAATGGGTCGTTACAGTGCAGAATTTG<br>CATTTAGATTTGTGAGCTCAGATTGTAGTTAAAAACACCCT<br>TGTTTCTACT</td><td>Prototype SHINE H5</td><td>|A/mute_swan/England/328136/2022|2022-10-03|2198908</td></tr>
<tr><td>H5_strain2_part</td><td>gBlocks Gene Fragment</td><td>GAAATTAATACGACTCACTATAGGGATCCCAAGTAA<br>ACGGGCAACGTGGAAGAATGGACTTCTTCTGGACAA<br>TTTTAAAACCAGATGATGCAATCCATTTCGAGAGTA<br>ATGGAAATTTCATTGCTCCAGAATATGCATATAAAA</td><td>SHINE-H5, SHINE-H5-LFA</td><td>|A/mute_swan/England/328136/2022|2022-10-0</td></tr>
</table>

| | | | | |
|---|---|---|---|---|
| | | TTGTCAAGAAAGGGGACTCAACAATTATGAAAAGTGGAGTGGAATATGGCCACTGCAACACCAAATGTCAAACCCCAGTAGGAGCGATAAATTCTAGTATGCCATTCCACAACATACATCCTCTCACCATTGGGGAATGCCCCAAATACGTGAAGTCAAACAAGTTGGTCCTTGCGACTGGGCTCAGAAATAGTCCTCTAAGAGAAAAGAGAAGAAAAAGAGGCCTGTTTGGGGCGATAGCAGGGTTTATAGAGGGAGGATGGCAGGGAATGGTTGATGGTTGGTATGGGTACCATCATAGCAATGAGCAGGGGAGTGGGTATGCTGCAGACAAAGAATCTACCCAAAA | | 3\|2198908 |
| H5_2.3.4.4b_strain1 | gBlocks Gene Fragment | GAAATTAATACGACTCACTATAGGGAACCAGAGGTTGGCACCAAAAATAGCTACTAGATCCCAAGTAAACGGGCAACGTGGAAGAATGGACTTCTTCTGGACAATCTTAAAACCAGATGATGCAATCCATTTCGAGAGTAACGGAAATTTCATTGCTCCAGAGTATGCATACAAAATTGTTAAGAAAGGGGACTCGACAATTATGAAAAGTGGAGTGGAATATGGCCACTGCAACACCAAATGTCAAACCCCAGTAGGTGCGATAAATTCTAGTATGCCATTTCACAACATACATCCTCTCACCATTGGGGAATGCCCCAAATACGTGAAATCAAACAAGTTGGTCCTTGCGACTGGGCTCAGAAATAGTCCTCTAAGAGAAAAGAGAAGAAAAAGAGGTCTGTTTGGGGCGAAGCAGGGTTTATAGAGGGAGGATGGCAGGGAATGGTTGATGGTTGGTATGGGTACCATCATAGCAATGAGCAGGGGAGTGGG | Prototype 2.3.4.4b clade-specific H5 SHINE, SHINE-H5-CS | \|A/Texas/37/2024\|2024-03-28\|EPI3171488 |
| H5_2.3.4.4b_strain2 | gBlocks Gene Fragment | GAAATTAATACGACTCACTATAGGGCACCAAAAATAGCTACTAGATCCCAAGTAAACGGGCAACGTGGAAGAATGGACTTCTTCTGGACAATCTTAAAACCAGATGATGCAATCCATTTCGAGAGTAACGGAAATTTCATTGCTCCAGAATATGCATACAAAATTGTTAAGAAAGGGGACTCGACAATTATGAAAAGTGGAGTGGAATATGGCCATTGCAACACCAAATGTCAAACCCCAGTAGGTGCGATAAATTCTAGTATGCCATTTCACAACATACATCCTCTCACCATTGGGGAATGCCCCAAATACGTGAAATCAAACAAGTTGGTCCTTGCGACTGGGCTCAGAAATAGTCCTCTAAGAGAAAAGAGAAGAAAAAGAGGTCTGTTTGGGGCGATAGCAGGGTTTATAGAGGGAGGATGGCAGGGAATGGTTGATGGTTGGTATGGGTACCATCATAGCAATGAGCAGGGGAGTGGGTACGCTGCGG | Prototype 2.3.4.4b clade-specific H5 SHINE, SHINE-H5-CS | \|A/dairy_cattle/Texas/2024\|2024-03-20\|EPI3158678 |
| H5_non2.3.4.4b_strain | gBlocks Gene Fragment | GAAATTAATACGACTCACTATAGGGCAAAAATAGCTACTAGATCCAAGGTAAACGGGCAAAGTGGAAGGATGGATTTCTTCTGGACAATTTTAAAACCGAATGATGCAATCAACTTCGAAAGTAATGGAAATTTCATTGCTCCAGAATATGCATACAAAATTGTCAAGAAAGGGGACTCAGCAATTATGAAAAGTGAATTGGAATATGGTAACTGCAACACCAAGTGTCAAACTCCAGTGGGGGCGATAAACTCTAGTATGCCATTCCACAACATACATCCTCTCACCATCGGGGAATGCCCCAAATATGTGAAATCAAACAGATTAGTCCTTGCGACTGGGCTCAGAAATAGCCCTCAAAGAGAGAGAAGAAAAAAAAAGAGGACTATTTGGAGCTATAGCAGGTTTTATAGAGGGAGGATGGCAGGGAATGGTAGATGGTTGGTATGGAT | Prototype 2.3.4.4b clade-specific H5 SHINE, SHINE-H5-CS | \|A/duck/Yunnan/5948/2003\|2003-01-01\|136318 |

| | | | | |
|---|---|---|---|---|
| | | ACCACCATAGCAATGAGCAGGGGAGTGGGTACGCTGCAGACAAAG | | |
| H7Eurasian_strain | gBlocks Gene Fragment | GAAATTAATACGACTCACTATAGGGGTCCTATAATGCTGAACTCTTGGTGGCAATGGAGAATCAACACACAATTGACCTGGCAGACTCAGAAATGAACAAACTATACGAGCGAGTGAAAAGGCAACTGAGAGAGAATGCTGAAGAAGATGGCACTGGCTGTTTTGAAATATTCCACAAGTGTGATGACGACTGCATGGCCAGTATCAGAAACAACACTTATGATCACAGCAAATACAGGGAGGAGGCAATGCAAAACCGAATACAGATTAACCCGGTCAAACTAAGCAGCAGTTACAAAGATGTGATACTTTGGTTTAGCTTCGGGGCATCATGTTTCATACTTCTTGCCATTGCAATGGGCCTTGTCTTCATATGTGTGAAGAATGGAAACATGCGGTGCACTATTTGTATATAAGTTTGGAAAAAACACC | SHINE-H7-Eurasian | \|A/chicken/Italy/1670/2015(H7N2) |
| H7American_strain | gBlocks Gene Fragment | GAAATTAATACGACTCACTATAGGGGTCGTATAATGCTGAACTGCTGGTAGCTATGGAAAATCAGCACACAATAGATCTTGCAGACTCAGAAATGAGCAAACTTTACGAGCGTGTAAGGAAACAACTGAGGGAGAATGCTGAAGAGGATGGGACTGGATGCTTTGAGATATTCCATAAGTGTGATGATCAATGCATGGAGAGCATAAGGAACAACACCTATGACCATACCCAATACAGAGCAGAGTCATTGCAGAATAGAATACAGATAGACCCAGTGAAACTGAGTAGTGGATACAAAGACATAATCTTATGGTTTAGCTTCGGGGCATCATGTTTTCTTCTTCTAGCCATTGCAATGGGATTGGTCTTCATTTGCATAAAGAATGGAAACATGCAGTGCACTATTTGTATATAGTTTGAGAAAAA | SHINE-H7-Eurasian | \|A/mallard/New York/AH0081558/2016(H7N7) |

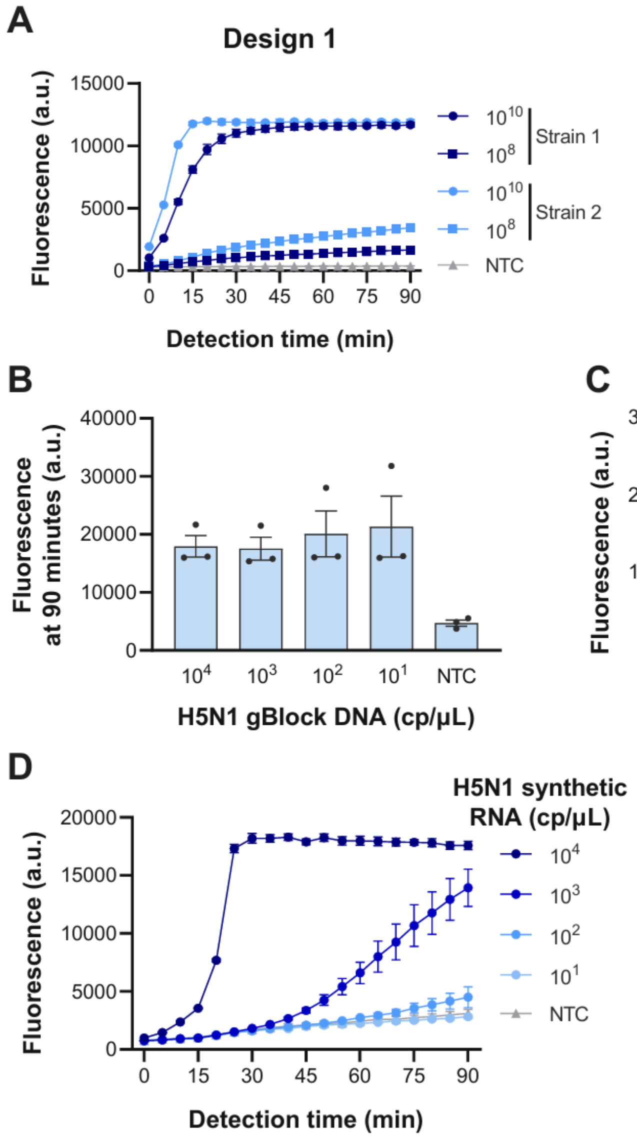


**Supplementary Figure 1. Kinetics of prototype H5 SHINE assay. A)** Kinetics from Figure 1C, separated by design. **B)** Fluorescence readout of H5N1 SHINE assays on a serial dilution of double-stranded DNA templates that encoded the entire H5 segment. **C)** Kinetics from Supplementary Figure 1B. **D)** Kinetics from Figure 1D. Timepoints represent mean fluorescence ± standard deviation of 3 technical replicates. NTC: no target control.

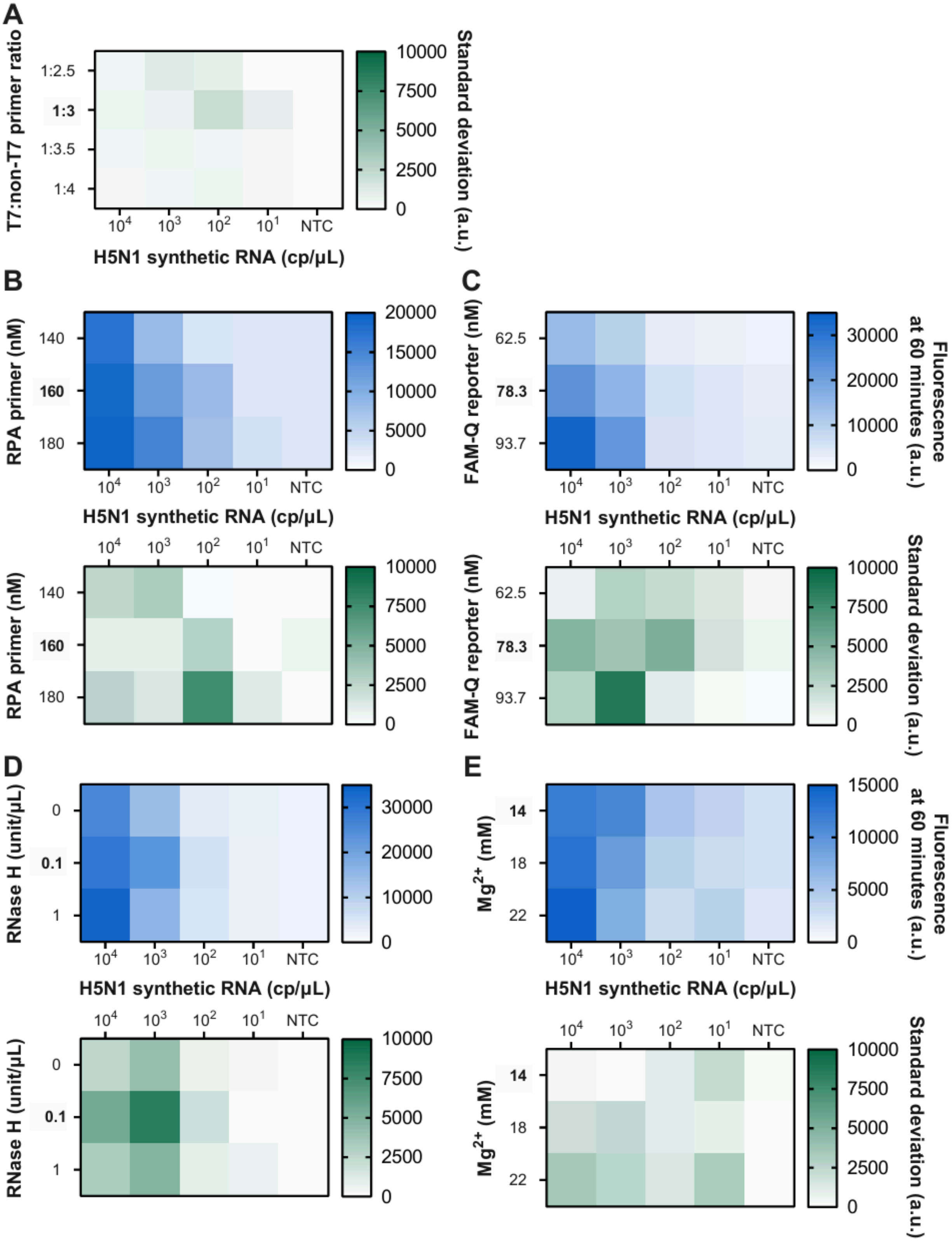
A
T7:non-T7 primer ratio
1:2.5
1:3
1:3.5
1:4
10⁴ 10³ 10² 10¹ NTC
H5N1 synthetic RNA (cp/μL)
10000 7500 5000 2500 0
Standard deviation (a.u.)
B
RPA primer (nM)
140
160
180
20000 15000 10000 5000 0
H5N1 synthetic RNA (cp/μL)
10000 7500 5000 2500 0
C
FAM-Q reporter (nM)
62.5
78.3
93.7
30000 20000 10000 0
Fluorescence at 60 minutes (a.u.)
Standard deviation (a.u.)
D
RNase H (unit/μL)
0
0.1
1
30000 20000 10000 0
E
Mg2+ (mM)
14
18
22
15000 10000 5000 0
NTC

**Supplementary Figure 2. Optimization of H5 SHINE assay (heatmap).** Heatmap showing the H5 SHINE assay performance at conditions with different **A)** RPA primer concentration, **B)** FAM-Q reporter concentration, **C)** RNase H concentration, **D)** $Mg^{2+}$ concentration. Blue represents mean fluorescence and green represents standard deviation of 3 technical replicates for each cell in heatmap. Bolded conditions (RPA primer at 160nM, FAM-Q reporter at 78.3nM, RNAse H at 0.1units/μL, $Mg^{2+}$ at 18mM) were selected for further development. NTC: no target control.

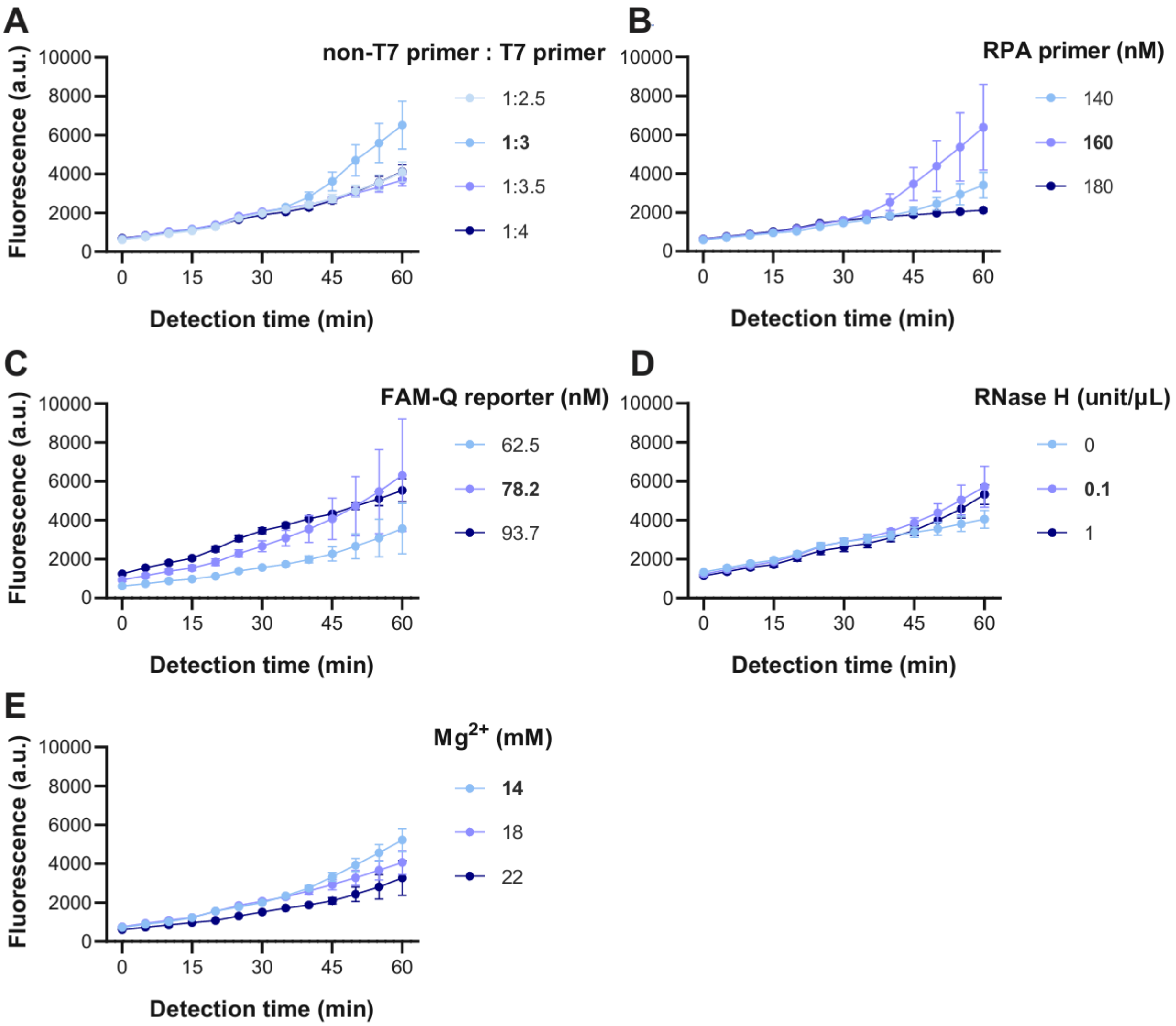


**Supplementary Figure 3. Kinetics of H5 SHINE assay optimization.** Kinetics from **A)** Figure 2A. **B)** Supplementary Figure 2B. **C)** Supplementary Figure 2C. **D)** Supplementary Figure 2D. **E)** Supplementary Figure 2E. All H5N1 synthetic RNA is at $10^2$ copies/μL. Timepoints represent mean fluorescence ± standard deviation of 3 technical replicates. Bolded conditions (non-T7 primer : T7 primer at 1:3 ratio, RPA primer at 160nM, FAM-Q reporter at 78.3nM, RNAse H at 0.1units/μL, $Mg^{2+}$ at 18mM) were selected for further development. Timepoints represent mean fluorescence ± standard deviation of 3 technical replicates. NTC: no target control.

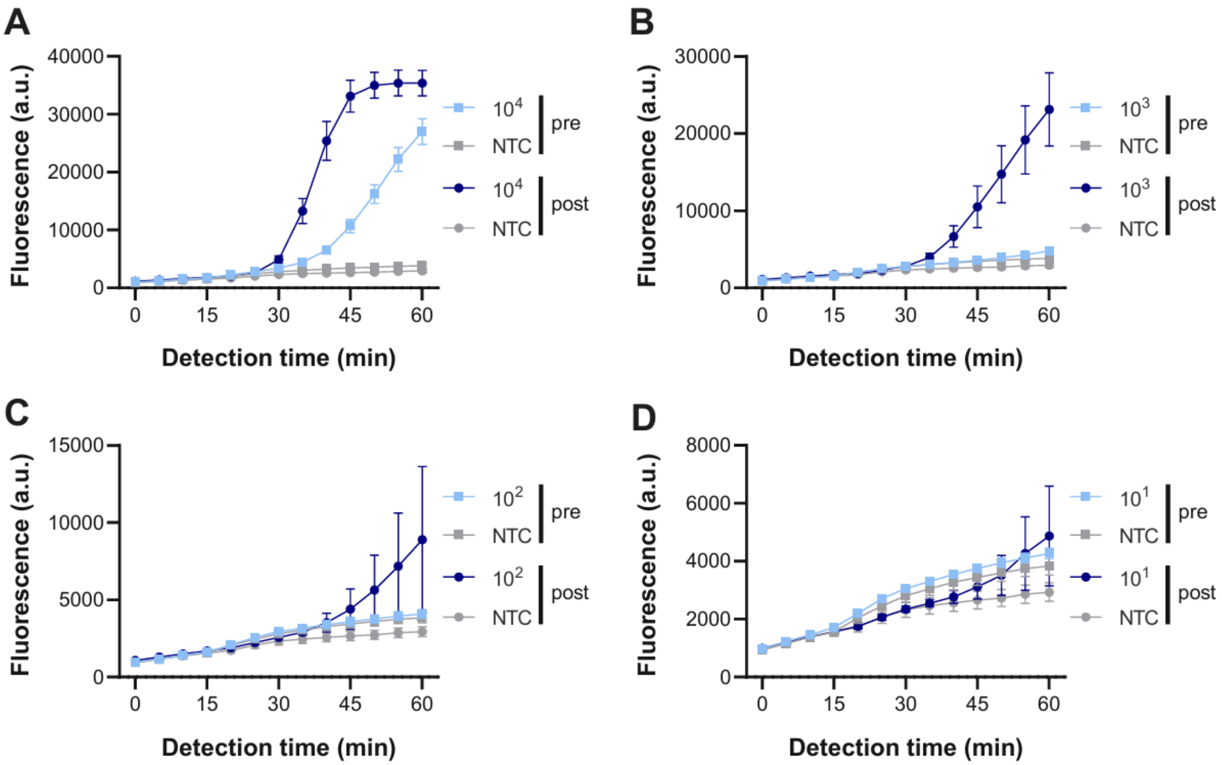


**Supplementary Figure 4. Kinetics of pre- vs post-optimization.** Kinetics from Figure 2B, separated by H5N1 synthetic RNA input concentration at **A)** $10^4$ cp/µL, **B)** $10^3$ cp/µL, **C)** $10^2$ cp/µL, **D)** $10^1$ cp/µL, Timepoints represent mean fluorescence ± standard deviation of 3 technical replicates. NTC: no target control, is shared in all panels.

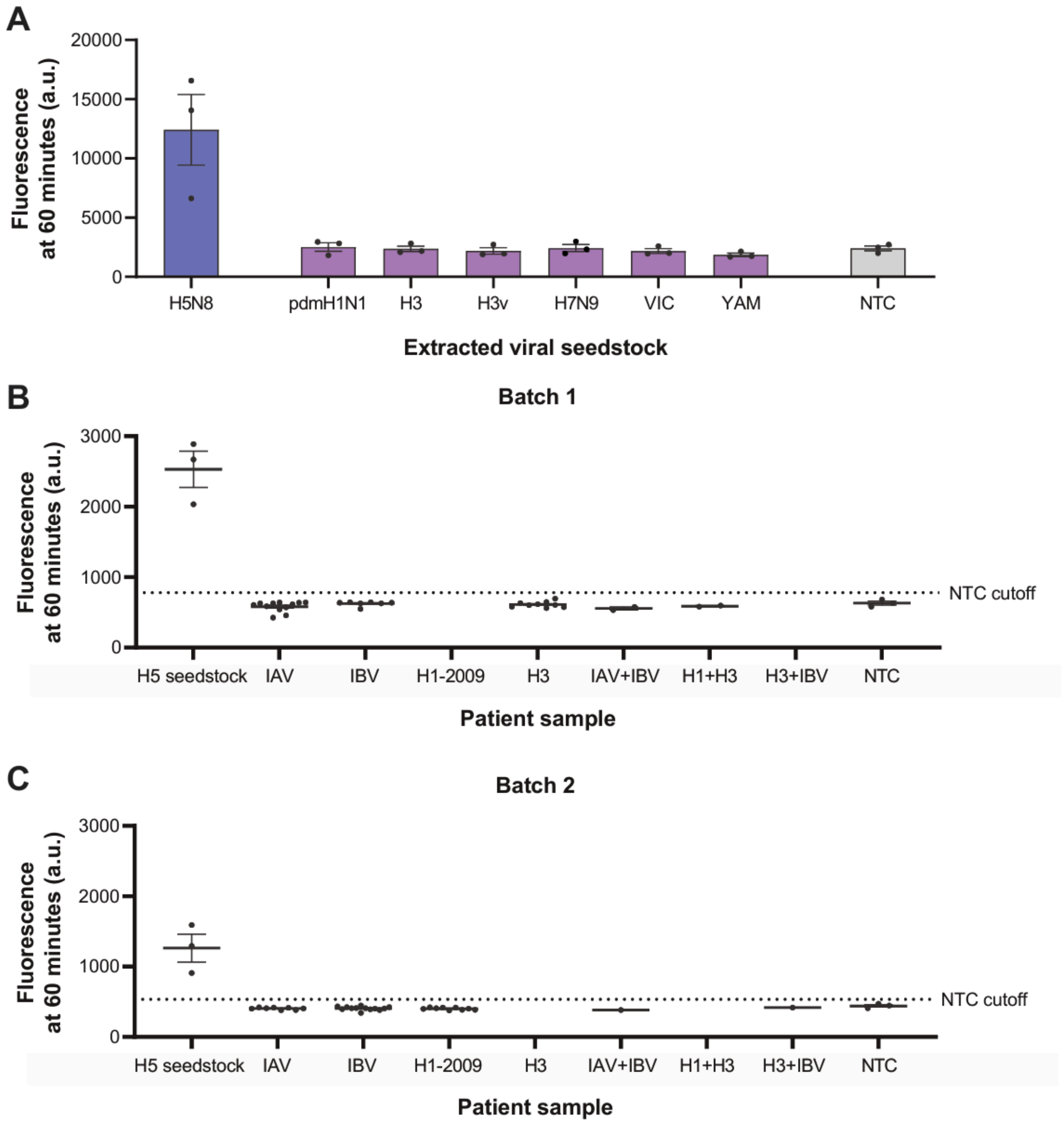


**Supplementary Figure 5. SHINE-H5 specificity test. A)** Cross-reactivity panel for SHINE-H5 against various seasonal influenza viral seedstocks. See Supplementary Table 2 for strain information. **B) C)** Fluorescence readout of optimized SHINE-H5 on RNA extracted from H5N1 viral seedstock ($10^2$ cp/µL) and from clinical NP swabs characterized by NAAT (BioFire) or qPCR. **B)** Batch 1. **C)** Batch 2. Each dot shows the mean of 3 technical replicates of a specific patient sample after 60 minutes. Cutoff is set as mean + three times standard deviation of 3 technical replicates of NTC (no target control). See Supplementary Table 3 for detailed patient information and qPCR/NAAT/SHINE readout.

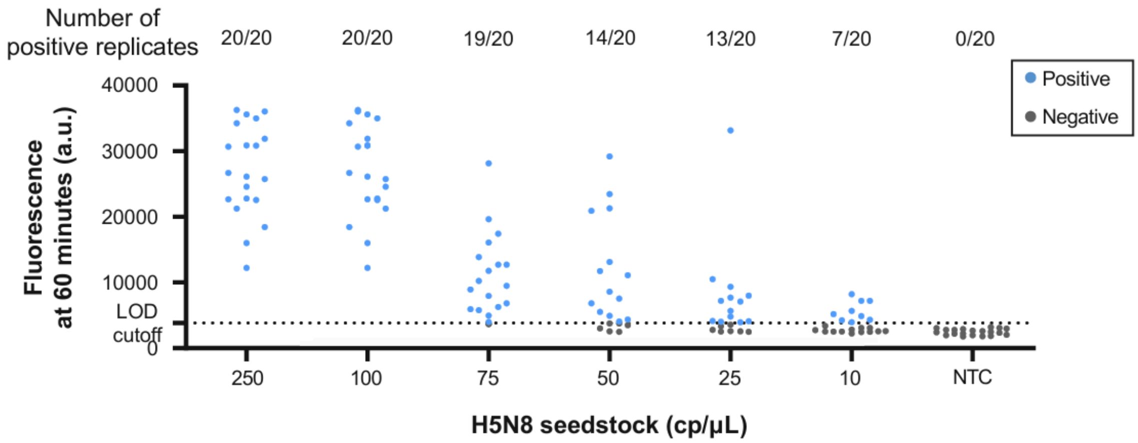


**Supplementary Figure 6. SHINE-H5 LOD.** Individual fluorescence values from Figure 2D. Cutoff is set as mean + three times standard deviation of 20 technical replicates of NTC (no target control).

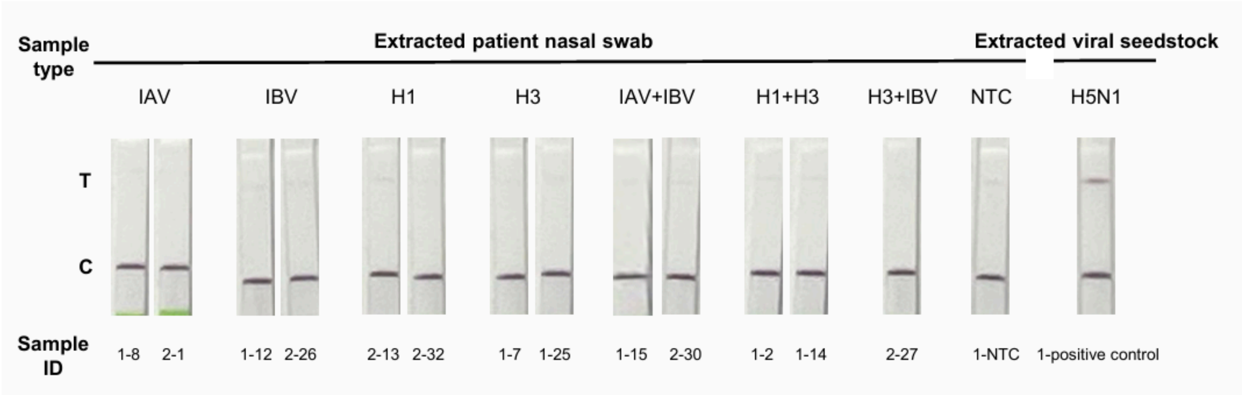


**Supplementary Figure 7. SHINE-H5-LFA specificity.** Paper-based colorimetric readout of SHINE-H5-LFA on RNA extracted from H5N1 viral seedstock ($10^2$ cp/µL) and from clinical NP swabs characterized by NAAT (BioFire) or qPCR after 60-minute incubation. NTC: no target control. See Supplementary Table 3 for detailed patient information and qPCR/NAAT/SHINE readout.

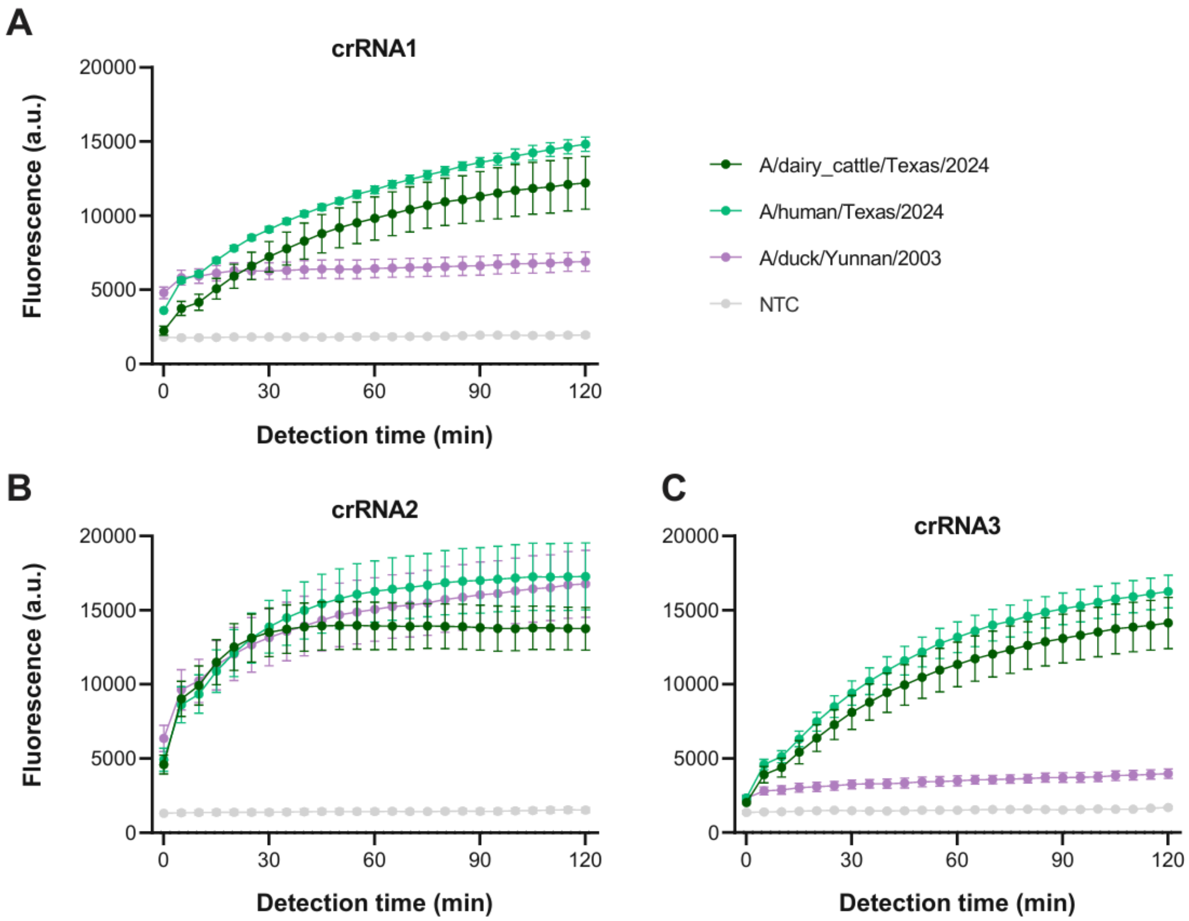


**Supplementary Figure 8. Kinetics of clade 2.3.4.4b A(H5N1) SHINE crRNA screening.** Kinetics from Figure 3C, separated by crRNA input **A)** crRNA 1, **B)** crRNA 2, **C)** crRNA 3. NTC: no target control.

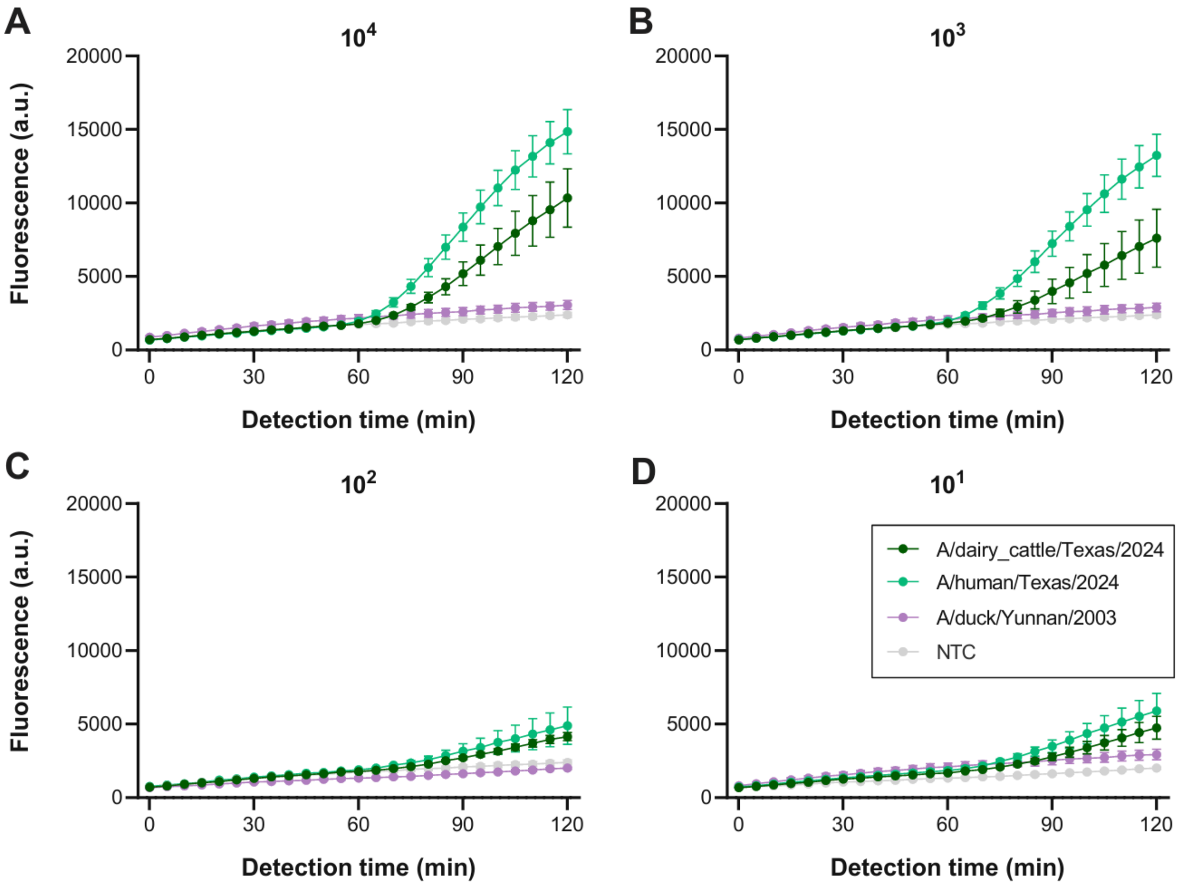


**Supplementary Figure 9. Kinetics of SHINE-H5-CS.** Kinetics from Figure 3D, separated by synthetic H5 RNA input concentration at **A)** $10^4$, **B)** $10^3$, **C)** $10^2$, **D)** $10^1$. Timepoints represent mean fluorescence ± standard deviation of 3 technical replicates. NTC: no target control.

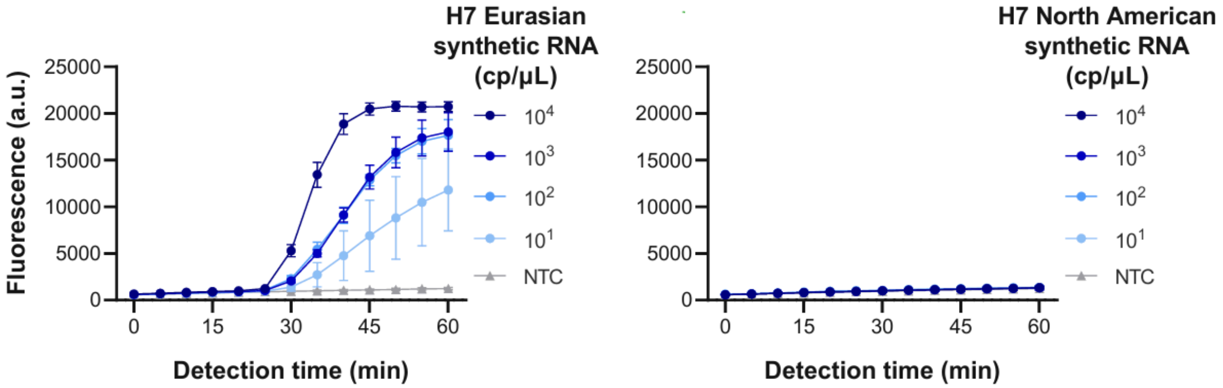


**Supplementary Figure 10. Kinetics of SHINE-H7-Eurasian.** Kinetics from Figure. 4C, separated by H7 RNA input type. Timepoints represent mean fluorescence ± standard deviation of 3 technical replicates. NTC: no target control.

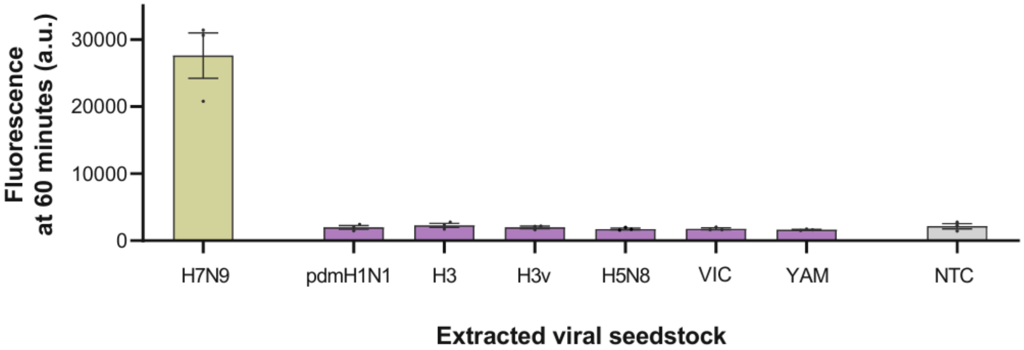


**Supplementary Figure 11. SHINE-H7-Eurasian specificity test.** Cross-reactivity panel for SHINE-H7-Eurasian against various seasonal influenza viral seedstocks. See Supplementary Table 3 for strain information.
